# Clinical Reference Percentiles for AI-derived Epicardial Adipose Tissue: A Multicenter Study

**DOI:** 10.64898/2026.08.28.26360111

**Authors:** Assiata Kamagate, Aakash Shanbhag, Mikolaj Buchwald, Robert J.H. Miller, Shaun Khanna, Tara Zuhair Kassem, Jacek Kwieciński, Renee Bullock-Palmer, Wenhao Zhang, Anna M Marcinkiewicz, Jirong Yi, Giselle Ramirez, Mark Lemley, Aditya Killekar, Paul B. Kavanagh, Joanna X. Liang, Leandro Slipczuk, Mark I. Travin, Erick Alexanderson, Isabel Carvajal-Juarez, René R.S. Packard, Mouaz Al-Mallah, Terrence D Ruddy, Robert A deKemp, Ronny R Buechel, Andrew J. Einstein, Wanda Acampa, Stacey Knight, Viet T Le, Steve Mason, Thomas L. Rosamond, Edward J Miller, Panithaya Chareonthaitawee, Daniel S. Berman, Damini Dey, Marcelo F. Di Carli, Piotr J Slomka

## Abstract

**Background and Aims:** Epicardial adipose tissue (EAT) has emerged as an important cardiovascular biomarker that reflects both inflammatory and cardiometabolic risk. EAT volume and density vary significantly across populations, yet there is a lack of multicenter studies investigating the predictive value of population-specific EAT percentiles.

**Methods:** In this multicenter study, we retrospectively analyzed low-dose computed tomography correction scans from 42,842 patients undergoing myocardial perfusion imaging. A derivation cohort of 15,082 patients was used to establish sex- and age-specific nomograms for EAT density and EAT volume indexed to body surface area. Percentile-based thresholds were tested for outcome prediction in a validation cohort of 27,760 patients. For clinical implementation, we developed an online EAT percentile calculator.

**Results:** Percentile curves demonstrated increased BSA-indexed EAT volume and decreasing EAT density with age. Over a median follow-up of 3.6 years (IQR: 1.83 – 5.14), 4,956 patients experienced a nonfatal myocardial infarction or death. In multivariable Cox models, patients above the 95^th^ sex- and age-specific percentile had significantly worse outcomes for BSA-indexed EAT volume [adjusted hazard ratio 1.30, 95% CI: 1.14 – 1.49, p < 0.001] and EAT density [adjusted hazard ratio 1.7, 95% CI: 1.51 – 1.92, p<0.001] when compared to patients below the 50^th^ percentile (p<0.001).

**Conclusion:** Age- and sex-specific EAT percentiles provide a clinically interpretable framework for contextualizing automated EAT measurements and identifying patients at increased cardiovascular risk. EAT density was a stronger prognostic marker and identified elevated risk even among patients with normal BMI, supporting its potential to provide information beyond conventional anthropometric assessment.

**Graphical Abstract:** 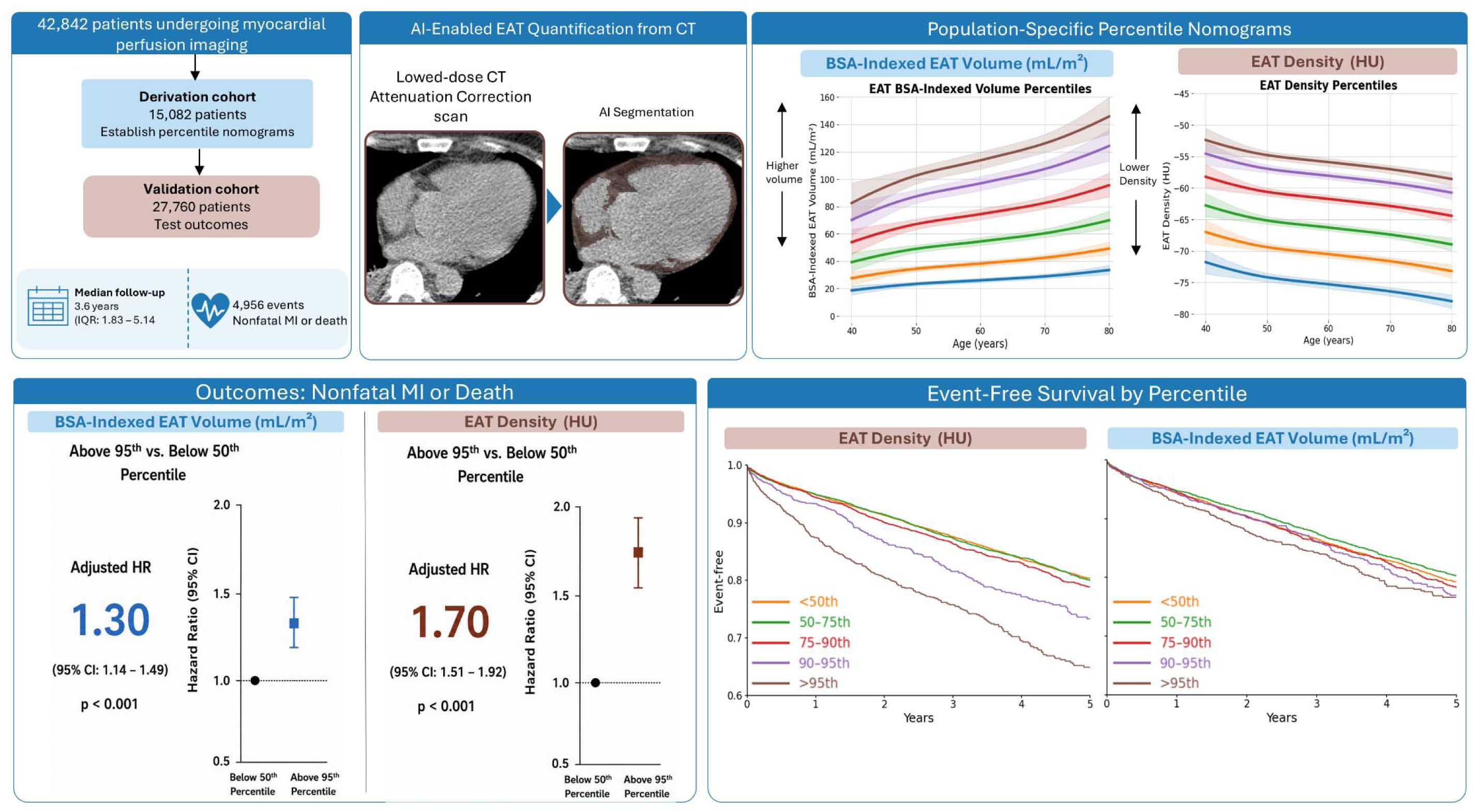

## INTRODUCTION

Epicardial adipose tissue (EAT) is a metabolically active fat depot located between the myocardium and visceral pericardium. Under physiological conditions, EAT exhibits cardioprotective properties, including fatty acid metabolism and secretion of anti-inflammatory adipokines [1, 2]. However, under pathological conditions including cardiometabolic disease, EAT undergoes functional changes and increases its release of proinflammatory adipokines, including tumor necrosis factor-alpha (TNF-α), interleukin-6 (IL-6), and leptin[3]. As a result, EAT may serve as a marker of underlying processes driving progression of coronary artery disease (CAD), the leading cause of death worldwide [4, 5].

Although body mass index (BMI) is a traditional marker of overall adiposity, it does not capture the fat distribution or biological activity of adipose tissue, such as its local paracrine and inflammatory effects [6, 7]. EAT volume and density may provide prognostic information beyond BMI by reflecting the distribution and burden of ectopic adipose tissues [8, 9]. In this regard, EAT volume is more closely associated with metabolic abnormalities, whereas EAT density is thought to reflect the local inflammatory state[10]. However, manual annotation of EAT density and volume can be time consuming, taking approximately 15 minutes per case [11]. Artificial intelligence (AI) can substantially reduce EAT quantification time (<2 seconds per case) and provides prognostic cardiovascular risk stratification with excellent clinical agreement [12, 13].

Even if density and EAT volume can be accurately estimated with AI methods, they vary significantly by age, sex, and race, complicating clinical decision making and underscoring the need for population-specific characterization [14–17]. Previous studies have examined these differences with binary thresholding; however, such methods do not capture the continuous and progressive relationship between EAT measures and cardiovascular risk [15, 18]. To date, there is a lack of large-scale multicenter studies evaluating population-specific EAT percentiles and their association with cardiovascular outcomes within a clinically interpretable framework. Percentile characterization of these key biomarkers may facilitate physician understanding and provide a mechanism for explaining results to patients. Therefore, in this study we aimed to leverage a unique multicenter myocardial perfusion imaging (MPI) dataset including over 40,000 low dose CT studies to determine sex-and age-specific, along with sex-, race-, and age-specific percentile thresholds of EAT volume and density and to test their predictive value.

## METHODS

### Study population

We considered data of 46,689 patients who underwent hybrid single-photon emission computerized tomography (SPECT)-CT MPI or positron emission tomography (PET)-CT MPI for evaluation of suspected or known coronary artery disease (CAD) from 2007 to 2024. Patients missing CT attenuation correction scans (CTAC), medical history, height, and weight were excluded from analysis (**Supplemental Figure 1**). An additional 500 patients used for the training of the AI EAT model in the previous work were also excluded from analysis[12]. Overall, the final study population (N=42,842) included 10,477 patients were from the international REgistry of Fast Myocardial Perfusion Imaging with NExt generation single proton emission computed tomography (REFINE SPECT) and 32,365 patients from the REgistry of Flow and Perfusion Imaging for Artificial Intelligence with positron emission tomography (REFINE PET) [19, 20]. The study was split into a derivation cohort comprised of 15,082 patients from 4 sites including Brigham & Women’s Hospital (PET), Ottawa Heart Institute (SPECT & PET), Houston Methodist (PET), and West Los Angeles Veterans Affair (PET) to establish population-specific percentiles. Derivation sites were selected to provide a cohort with age, sex, and racial distribution representative of the overall study population. A validation cohort of 27,760 patients from the remaining 11 sites were utilized to validate percentile cutoffs on an adjusted Cox model (**Supplemental Table 1**). Images were reconstructed with attenuation correction for both SPECT and PET CT MPI [19, 20]. All imaging files were de-identified at each site prior to transfer to the core laboratory and underwent quality control by experienced technologists at the core laboratory (Cedars-Sinai Medical Center, Los Angeles, CA). Additional details on image acquisition parameters for CTAC scans for each site are available in **Supplemental Table 2.** The study complied with the Declaration of Helsinki and received Institutional Review Board approval from Cedars-Sinai Medical Center for the overall study. Sites obtained either written informed consent or a waiver of consent for the use of deidentified data [19, 20].

### Percentile Stratification

Sex-and age-specific, along with sex-, race-, and age-specific percentile thresholds were constructed using a derivation cohort. We fit a regression model to predict EAT density (HU) and BSA-indexed EAT volume (mL/m²) as a function of age. EAT volume was specifically modeled on a log-scale due to the skewed nature of the data, especially at extreme values. Residuals for each patient were computed as the difference between the observed and model predicted EAT value. We then pooled the residuals and calculated the empirical residual percentiles (10^th^, 25^th^, 50^th^,75^th^,90^th^, 95^th^) [21, 22]. To obtain age-specific percentiles, we fit a model to predict the expected EAT density or BSA-indexed EAT volume for each observed age integer within sex and race subgroups. We then added the residual percentile offsets to predicted EAT density and BSA-indexed EAT volume at each age **(Supplemental Figure 2)**. We summarized the age-specific percentiles curves in defined age categories: <45, 45-54, 55-64, 65-74, and ≥75 years by averaging the predicted percentile values across age within each category.

Sex- and race-specific percentile cutoffs were obtained (separately for EAT density and BSA-indexed EAT volume) and then applied to the validation cohort to classify each patient into one of five percentile groups: <50^th^, 50-75^th^, 75^th^-90^th^, 90-95^th^, and >95^th^.

Confidence intervals were estimated using nonparametric bootstrap resampling (B=1000). Individuals were resampled with replacement within sex and race subgroups, the age model was refit, residual percentiles were recomputed, and age-specific percentile curves were reconstructed and summarized into 5 age bins. For each sex-and age-specific and race-and age-specific percentile, 95% confidence intervals were derived from the 2.5^th^ and 97.5^th^ percentiles of the bootstrap distribution.

### EAT Calculator

To facilitate translation into clinical practice, developed a freely accessible online calculator that provides individualized age- and sex-specific EAT percentiles [23].

### Clinical data

Baseline characteristics data in this study included patients’ age, sex, race, BMI, BSA, smoking status, hypertension, diabetes, dyslipidemia, prior CAD, past myocardial infarction (MI), family history of CAD, and coronary artery calcium (CAC) scores. Race in this study was defined as a self-reported construct; patients racially identifying as White or identifying as Black or African American were used as a sub-cohort for analysis. Other self-reported races were not assessed due to an insufficient sample size or indicating their race as unknown for race-specific analyses.

### Outcomes

The primary clinical outcome was all-cause mortality or non-fatal MI. Non-fatal MI was defined as hospital admission for new or worsening chest pain with elevated cardiac enzyme levels and ischemic ECG changes [19]. All-cause mortality was determined by the retrieval of National Death Index (in the United States), hospital electronic health records, physician offices, or medical chart reviews. Patients who experienced both death and MI were assigned the earliest event interval. All events were adjudicated by experienced physicians at each site [20].

### Calculating EAT amounts

The volume and density of EAT were measured using the previously published EAT segmentation method [12]. The model first identifies the silhouette of the pericardial sac, and then automatically segments EAT based on attenuation thresholds (-190 Hounsfield units [HU] to –30 HU). The superior and inferior boundaries of the pericardium were identified as the bifurcation of the pulmonary trunk and the posterior descending artery, respectively. A median filter of radius 3 voxels is used in the model to reduce the speckle noise [12]. Automatic quality control for epicardial contour masks was also carried out as previously described in [24]. When EAT volume is used for risk stratification, it is first indexed by the patient’s body surface area (BSA) parameter [9].

### Statistical analysis

Baseline characteristics were summarized separately by derivation and validation cohort. Categorical variables were summarized as number, frequency and percentage and compared with Pearson’s Chi-squared test, while continuous variables were summarized as median and interquartile range and were compared with Wilcoxon rank sum test.

Cox proportional hazard models were used to find the association between EAT density and BSA-indexed EAT volume percentiles and the outcome of death or MI. We applied population stratified thresholds to the derivation cohort, with patients below the 50^th^ percentile for their sex-and age and their race-and sex serving as the reference group. The model was adjusted for site, dyslipidemia, diabetes mellitus, smoking, previous CAD, family history of CAD, BMI hypertension, past MI, stress total perfusion deficit (TPD), stress ejection fraction (EF), and CAC score. Imaging modality was incorporated as a frailty term to account for differences in imaging modality. To account for differences in imaging parameters, supplemental analysis included additional adjustment for tube current and tube voltage. Sex-and age-specific percentiles were additionally adjusted for race. To determine whether EAT percentiles provided prognostic information beyond generalized adiposity, analyses were repeated within normal-weight, overweight, and obese BMI categories. To evaluate whether EAT percentiles retain prognostic value in a lower risk-risk population, analyses were repeated in patients with a myocardial flow reserve greater than 2 and a CAC of 0. Missing stress ejection fraction values were imputed using the cohort median prior to Cox proportional hazards modeling. Additional details on missing variables are shown in **Supplemental Table 3**.

Kaplan – Meier curves were used to estimate the event-free survival probabilities across EAT density and BSA-indexed EAT volume percentile groups, with death or MI as the outcome. Group differences were compared using the log-rank test. P values <0.05 were considered significant. Analyses were performed using R software version 4.1.2 (R foundation for statistical computing), and Python version 3.11.5 (Python software foundation).

## RESULTS

### Patient characteristics

In total, 15,082 patients from 4 sites were included in the derivation cohort with median age 67 (IQR 59,75) and 58% male. The validation cohort consisted of 27,760 patients from 11 sites with median age 67 (IQR 58,75) and 58% male. Most patients self-reported their race as White in both the derivation (68%) and validation cohort (66%). Additional details on the derivation and validation cohorts are shown in **Table 1**. Median EAT density was higher in women than men [-66 (IQR -71, -61) vs -67 (IQR -72, -62) HU, p<0.001]. BSA-indexed EAT volume was higher in men than women [61.1 (IQR 42.3, 83.1) vs 52.8 (IQR 35.8, 74.3) mL/m², p<0.001]. Additional details on demographic and clinical differences between men and women can be found in **Supplemental Table 4.**

**Table 1:** Patient characteristics by cohort. Categorical variables are shown as n (%) and were compared with Pearson’s Chi-squared test, while continuous variables are shown as median Interquartile range (IQR) and were compared with Wilcoxon rank sum test. Abbreviations: BMI – body mass index, BSA – body surface area, CAD – coronary artery disease, EAT – epicardial adipose tissue, HU – Hounsfield unit

| Characteristic | Overall<br>N = 42,842 | Derivation<br>N = 15,082 | Validation<br>N = 27,760 | P value |
| --- | --- | --- | --- | --- |
| Male | 24,889 (58%) | 8,722 (58%) | 16,167 (58%) | 0.4 |
| Age | 67.0 (58.0, 75.0) | 67.0 (59.0, 75.0) | 67.0 (58.0, 75.0) | 0.002 |
| Race |  |  |  | <0.001 |
| *American Indian or Alaska Native* | 157 (0.4%) | 68 (0.5%) | 89 (0.3%) |  |
| *Asian* | 1,147 (2.7%) | 466 (3.1%) | 681 (2.5%) |  |
| *Black or African American* | 4,640 (11%) | 1,659 (11%) | 2,981 (11%) |  |
| *Native Hawaiian or Other Pacific Islander* | 199 (0.5%) | 35 (0.2%) | 164 (0.6%) |  |
| *White* | 28,633 (67%) | 10,315 (68%) | 18,318 (66%) |  |
| *Unknown* | 8,066 (19%) | 2,539 (17%) | 5,527 (20%) |  |
| BMI | 29.4 (25.5, 34.6) | 29.8 (25.8, 34.8) | 29.1 (25.3, 34.4) | <0.001 |
| BSA (m<sup>2</sup>) | 2 (1.8, 2.2) | 2 (1.8, 2.2) | 2 (1.8, 2.2) | <0.001 |
| Hypertension | 31,747 (74%) | 11,511 (76%) | 20,236 (73%) | <0.001 |
| Dyslipidemia | 29,022 (68%) | 11,251 (75%) | 17,771 (64%) | <0.001 |
| Diabetes Mellitus | 14,499 (34%) | 5,301 (35%) | 9,198 (33%) | <0.001 |
| Family History of CAD | 11,940 (28%) | 3,246 (22%) | 8,694 (31%) | <0.001 |
| Past Myocardial Infarction | 7,131 (17%) | 3,095 (21%) | 4,036 (15%) | <0.001 |
| Smoking | 10,027 (23%) | 4,478 (30%) | 5,549 (20%) | <0.001 |
| CAC score | 139.6 (0, 977.9) | 137.8 (0, 988.7) | 140.7 (0, 974.3) | 0.015 |
| Left ventricle volume (mL) | 122.5 (100.1, 151.2) | 124.8 (101.4, 153.9) | 121.1 (99.2, 149.6) | <0.001 |
| EAT volume (mL) | 112.6 (74.6, 161.2) | 109.9 (72.0, 157.7) | 114.1 (76.0, 163.2) | <0.001 |
| EAT indexed BSA (mL/m<sup>2</sup>) | 57.4 (39.3, 79.7) | 55.8 (38.0, 77.5) | 58.4 (40.1, 80.9) | <0.001 |
| EAT % heart size | 19.5 (13.1, 27.7) | 18.3 (12.3, 25.7) | 20.2 (13.6, 28.7) |  |
| EAT density (HU) | -67.0 (-71.0, -62.0) | -66.0 (-71.0, -62.0) | -67.0 (-71.0, -62.0) | <0.001 |

### EAT Population Distributions

BSA-indexed EAT volumes were higher in male patients and increased with age. At age 40, the 50^th^ percentile BSA-indexed EAT volume was 44.4 mL/m² in males and 36.1 mL/m² in females, increasing to 67.3 mL/m² and 61.2 mL/m² by age 80, respectively **(Supplemental Table 5)**. EAT density was higher in females than in males, with both groups demonstrating a decrease in density with age (**Figure 1)**. EAT median density, decreased from -64.1 HU to -68.6 in males, and -62.1 HU to -67.7 HU in females **(Supplemental Table 6).** An example of the EAT percentile calculator showcasing a female in her 60s is shown in **Supplemental Figure 3.**

**Figure 1.**
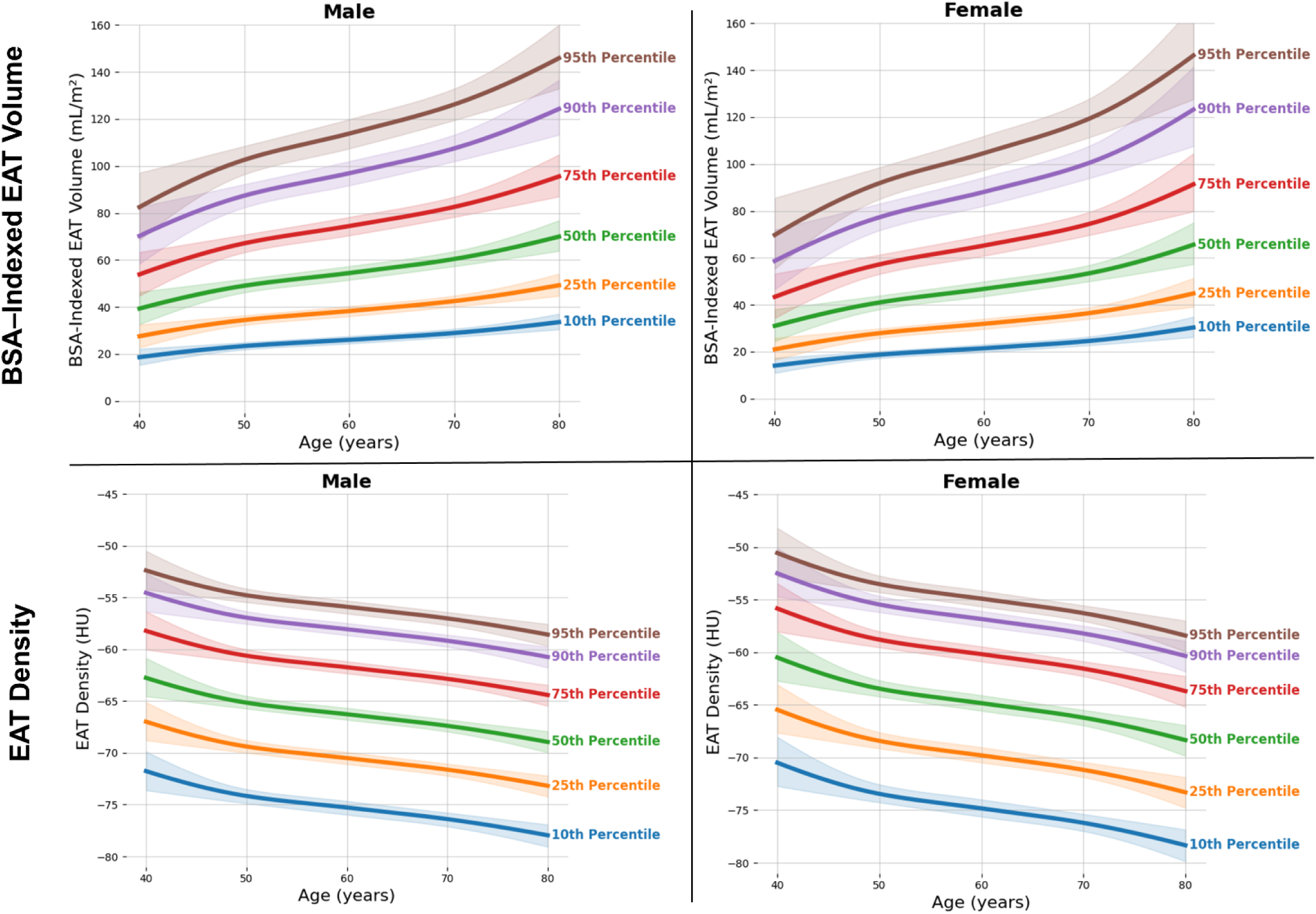
Sex- and age-specific percentiles for EAT density and BSA-indexed EAT volume with 95% confidence intervals

Among White patients, BSA-indexed EAT volume was consistently higher in males across all age groups. However, among Black patients, while 50^th^ percentile values remained higher in males, upper percentile values (≥90^th^) were higher in female patients, suggesting differences in distributional spread **(Supplemental Figure 4)**. Additionally, EAT density was higher in females and declined with age in both racial cohorts **(Supplemental Figure 5).**

### Clinical Outcomes

During a median follow-up of 3.6 years (IQR: 1.83 – 5.14), 4,956 patients in the validation cohort experienced a nonfatal MI or death. Patients who experienced death and MI had higher BSA-indexed EAT volumes than patients who did not have an event [59.2 (39.6, 83.3) vs 57 (39.3, 79) mL/m², p<0.001]. Additional event-stratified patient details are shown in **Table 2**.

**Table 2:** Characteristics of patients that experienced a death or MI event versus those that did not. Categorical variables are shown as n (%) and were compared with Pearson’s Chi-squared test, while continuous variables are shown as median Interquartile range (IQR) and were compared with Wilcoxon rank sum test. Abbreviations: MI – myocardial infarction, BMI – body mass index, BSA – body surface area, CAD – coronary artery disease, CAC – Coronary Artery Calcium, EAT – epicardial adipose tissue, HU – Hounsfield unit

| Characteristic | Overall<br>N = 42,842 | Death or MI<br>N = 8,361 | No Death or MI<br>N = 34,481 | P<br>value |
| --- | --- | --- | --- | --- |
| Male | 24,889 (58%) | 5,342 (64%) | 19,547 (57%) | <0.001 |
| Age | 67.0 (58.0, 75.0) | 71.0 (63.0, 79.0) | 66.0 (57.0, 74.0) | <0.001 |
| Race |  |  |  | <0.001 |
| *American Indian or*<br>*Alaska Native* | 157 (0.4%) | 43 (0.5%) | 114 (0.3%) |  |
| *Asian* | 1,147 (2.7%) | 188 (2.2%) | 959 (2.8%) |  |
| *Black or African*<br>*American* | 4,640 (11%) | 932 (11%) | 3,708 (11%) |  |
| *Native Hawaiian or Other*<br>*Pacific Islander* | 199 (0.5%) | 49 (0.6%) | 150 (0.4%) |  |
| *Unknown* | 8,066 (19%) | 948 (11%) | 7,118 (21%) |  |
| *White* | 28,633 (67%) | 6,201 (74%) | 22,432 (65%) |  |
| BMI | 29.4 (25.5, 34.6) | 28.2 (24.5, 33.3) | 29.7 (25.7, 34.8) | <0.001 |
| BSA (m<sup>2</sup>) | 2.0 (1.8, 2.2) | 1.9 (1.8, 2.1) | 2.0 (1.8, 2.2) | <0.001 |
| Hypertension | 31,747 (74%) | 6,969 (83%) | 24,778 (72%) | <0.001 |
| Dyslipidemia | 29,022 (68%) | 6,076 (73%) | 22,946 (67%) | <0.001 |
| Diabetes Mellitus | 14,499 (34%) | 3,675 (44%) | 10,824 (31%) | <0.001 |
| Family History of CAD | 11,940 (28%) | 2,145 (26%) | 9,795 (28%) | <0.001 |
| Past Myocardial Infarction | 7,131 (17%) | 2,583 (31%) | 4,548 (13%) | <0.001 |
| Smoking | 10,027 (23%) | 2,531 (30%) | 7,496 (22%) | <0.001 |
| CAC score | 139.6 (0.0, 977.9) | 525.2 (52.5, 1,719.6) | 89.6 (0.0, 784.0) | <0.001 |
| Left ventricle volume (mL) | 122.5 (100.1, 151.2) | 135.8 (108.7, 170.9) | 119.9 (98.4, 146.6) | <0.001 |
| EAT volume (mL) | 112.6 (74.6, 161.2) | 114.9 (73.6, 166.7) | 112.0 (74.8, 160.0) | 0.012 |
| EAT indexed BSA (mL/m<sup>2</sup>) | 57.4 (39.3, 79.7) | 59.2 (39.6, 83.3) | 57.0 (39.3, 79.0) | <0.001 |
| EAT % heart size | 19.5 (13.1, 27.7) | 18.0 (11.8, 25.9) | 19.9 (13.4, 28.0) |  |
| EAT density (HU) | -67 (-71, -62) | -66 (-71, -61) | -67 (-71, -62) | <0.001 |

In an adjusted Cox proportional hazards model, there was an increased risk of death or MI with increased EAT density and BSA-indexed EAT volume. Compared to the reference group (<50^th^ percentile), density outperformed volume, with patients above the 95^th^ EAT density percentile having a 70% higher risk of experiencing death or MI [adjusted hazard ratio 1.70, 95% CI: 1.51 – 1.92, p<0.001], while patients above the 95^th^ percentiles for BSA-indexed EAT volume had 30% increased risk [ adjusted hazard ratio 1.30, 95% CI: 1.14 – 1.49, p < 0.001] **(Figure 2)**. Moreover, after additionally adjusting the model for tube current and tube voltage, patients above the 95^th^ percentile for EAT density and BSA-indexed EAT volume were still at significantly higher risk of death or MI **(Supplemental Figure 6)**. Across BMI categories, higher EAT density percentiles remained significantly associated with adverse outcomes in obese patients, whereas elevated BSA-indexed EAT volume percentiles demonstrated the strongest prognostic value among normal-weight individuals **(Figure 3)**. EAT density percentiles retained strong prognostic value in a lower-risk population, with patients above the 95^th^ percentile having a 91% higher risk of experiencing an event [adjusted hazard ratio 1.91, 95% CI: 1.18 – 3.08, p<0.001]. Adjusted Cox models for BSA-indexed EAT volume are shown **in Supplemental Figure 7**. Unadjusted Cox proportional hazards models are shown in **Supplemental Figure 8** and race-stratified percentiles are shown in **Supplemental Figure 9 and 10** and follow similar patterns. Unadjusted race-stratified cox proportional hazards models are shown in **Supplemental Figure 11.**

**Figure 2.**
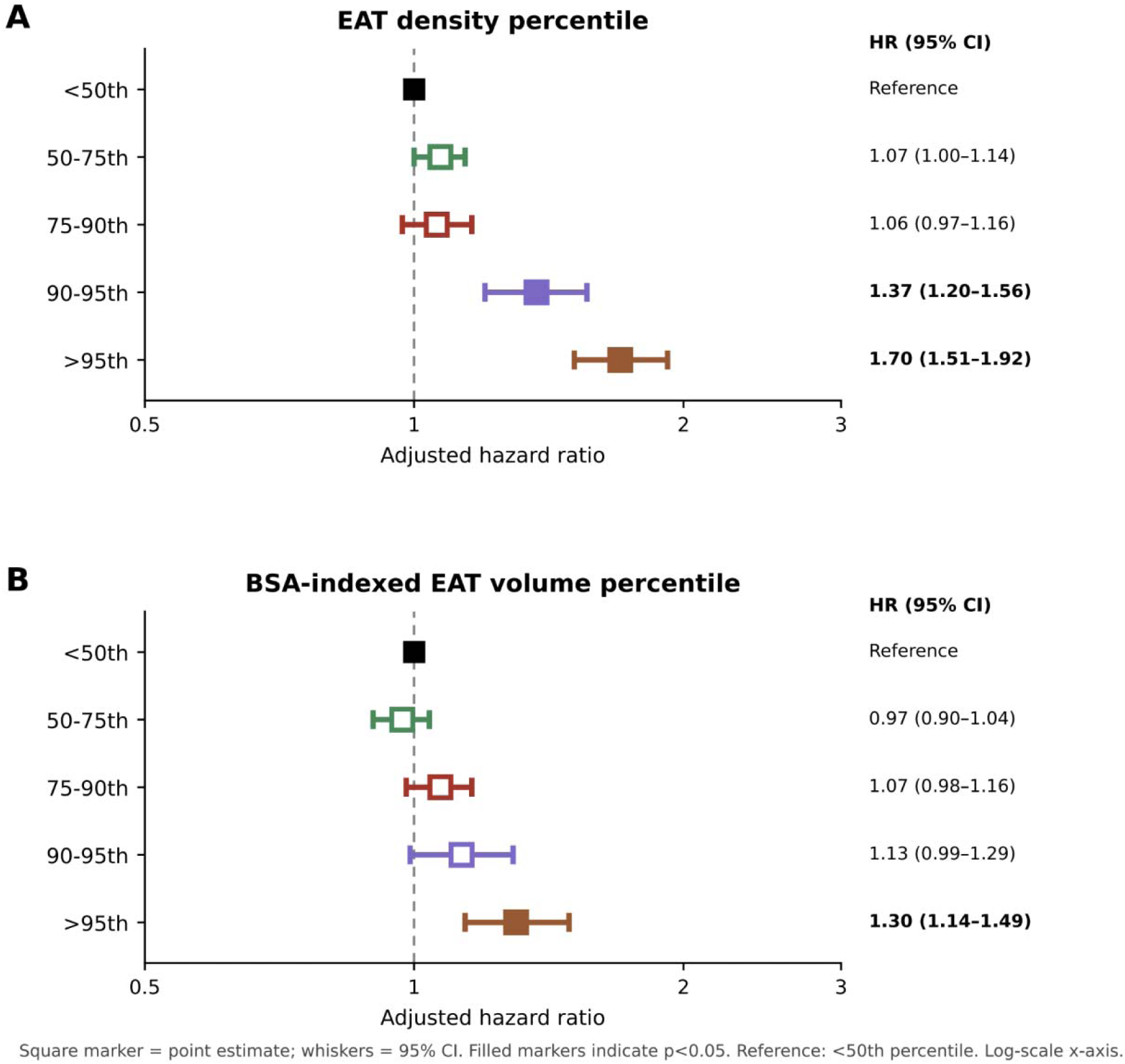
Adjusted Cox proportional hazards model of sex- and age-stratified EAT percentile groups, with patients below the 50^th^ percentile serving as the reference group. Percentile thresholds were derived in a derivation cohort and applied to a validation cohort; therefore, group sizes in the validation cohort may differ from the expected 50%, 25%, 15%, 5%, and 5% distribution. Panel A displays adjusted hazard ratios across EAT density percentiles. Panel B displays adjusted hazard ratios across BSA-indexed EAT percentiles. Models were adjusted for site, race, dyslipidemia, diabetes mellitus, smoking, previous coronary artery disease, family history of coronary artery disease, body mass index, hypertension, past myocardial infarction, coronary artery calcium score, stress total perfusion deficit, and stress ejection fraction, along with imaging modality as shared frailty. N = 27,760

**Figure 3.**
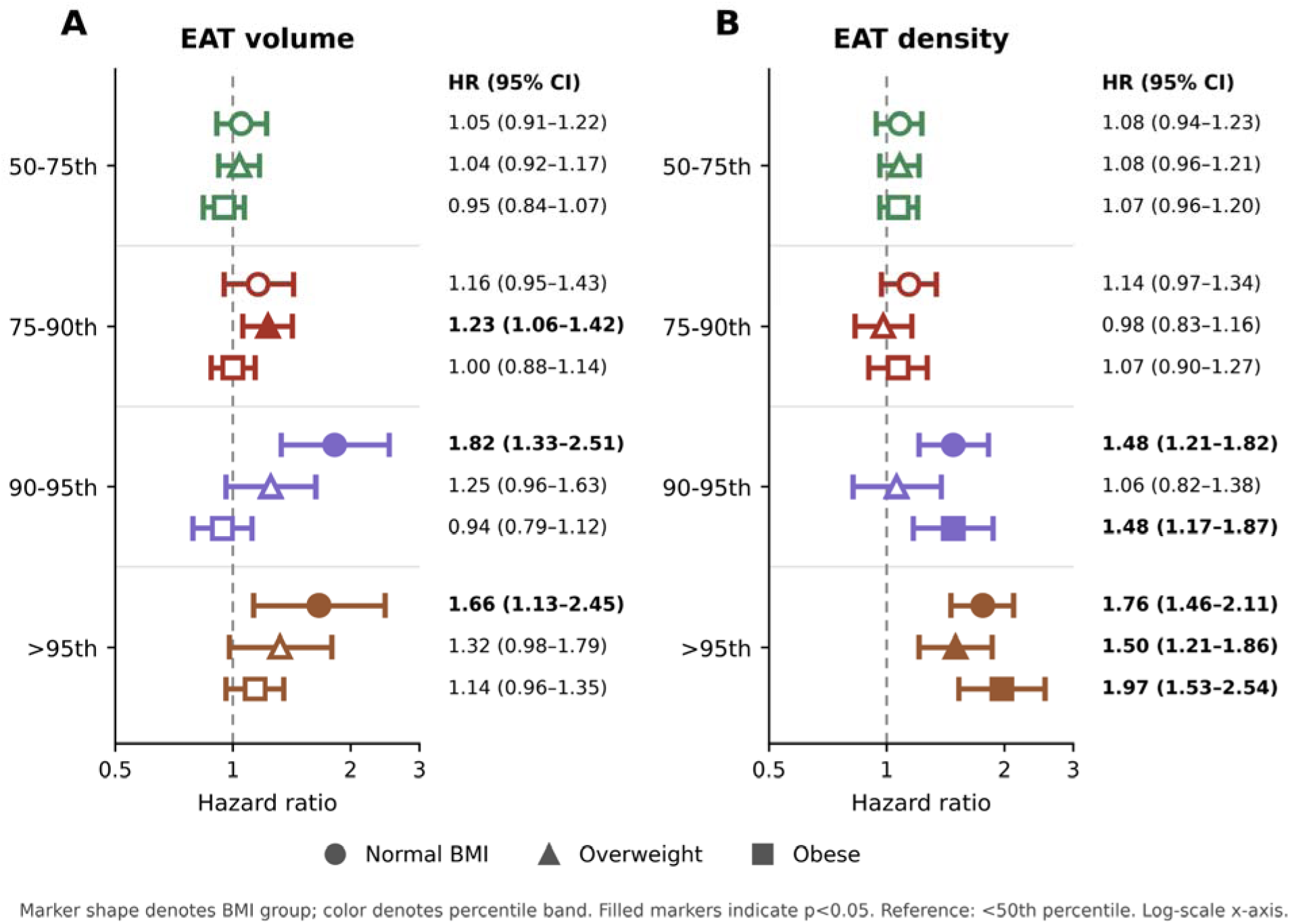
Adjusted Cox proportional hazards model of sex- and age-stratified EAT percentile groups in patients with Normal BMI, overweight patients, and obese patients. Models were adjusted for site, race, dyslipidemia, diabetes mellitus, smoking, previous coronary artery disease, family history of coronary artery disease, hypertension, past myocardial infarction, coronary artery calcium score, stress total perfusion deficit, and stress ejection fraction, along with imaging modality as shared frailty. N = 27,760

Kaplan-Meier curves stratified by sex- and age-demonstrate that patients in higher EAT density (log-rank p<0.001) and BSA-indexed EAT volume percentile groups (log-rank p<0.005) were more likely to experience death or MI (**Figure 4**). Additionally, Kaplan-Meier curves stratified by race-, sex-, and age-specific percentiles demonstrate that Black and White patients in higher EAT density and BSA-indexed EAT volume percentiles were more likely to experience death or MI (**Supplemental Figure 12)**.

**Figure 4.**
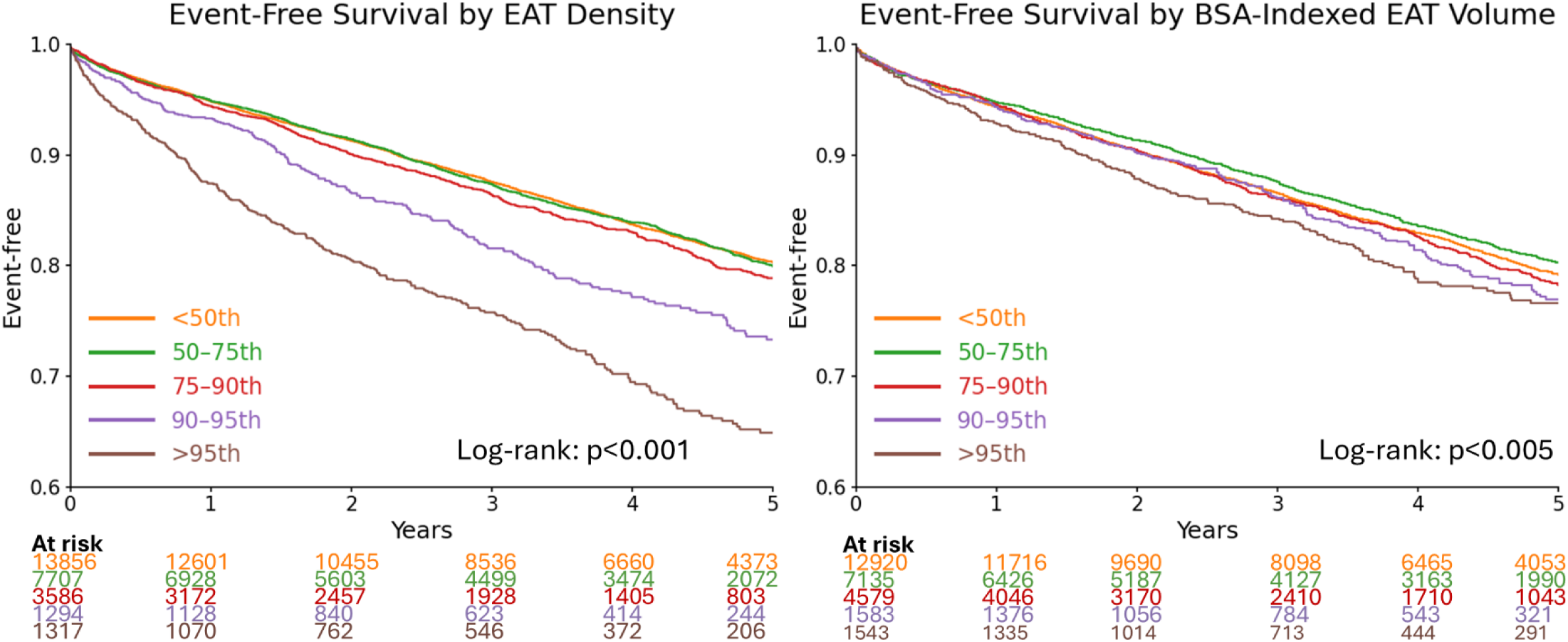
Event-free Kaplan –Meier curves of EAT Density and BSA-indexed EAT volume percentile groups. Percentile groups were stratified by sex and age. Kaplan –Meier curves were compared using log-rank test

### Case Examples

Four cases demonstrating patients in different EAT density and BSA-indexed volume percentiles for their age and sex are shown in **Figure 5**.

**Figure 5.**
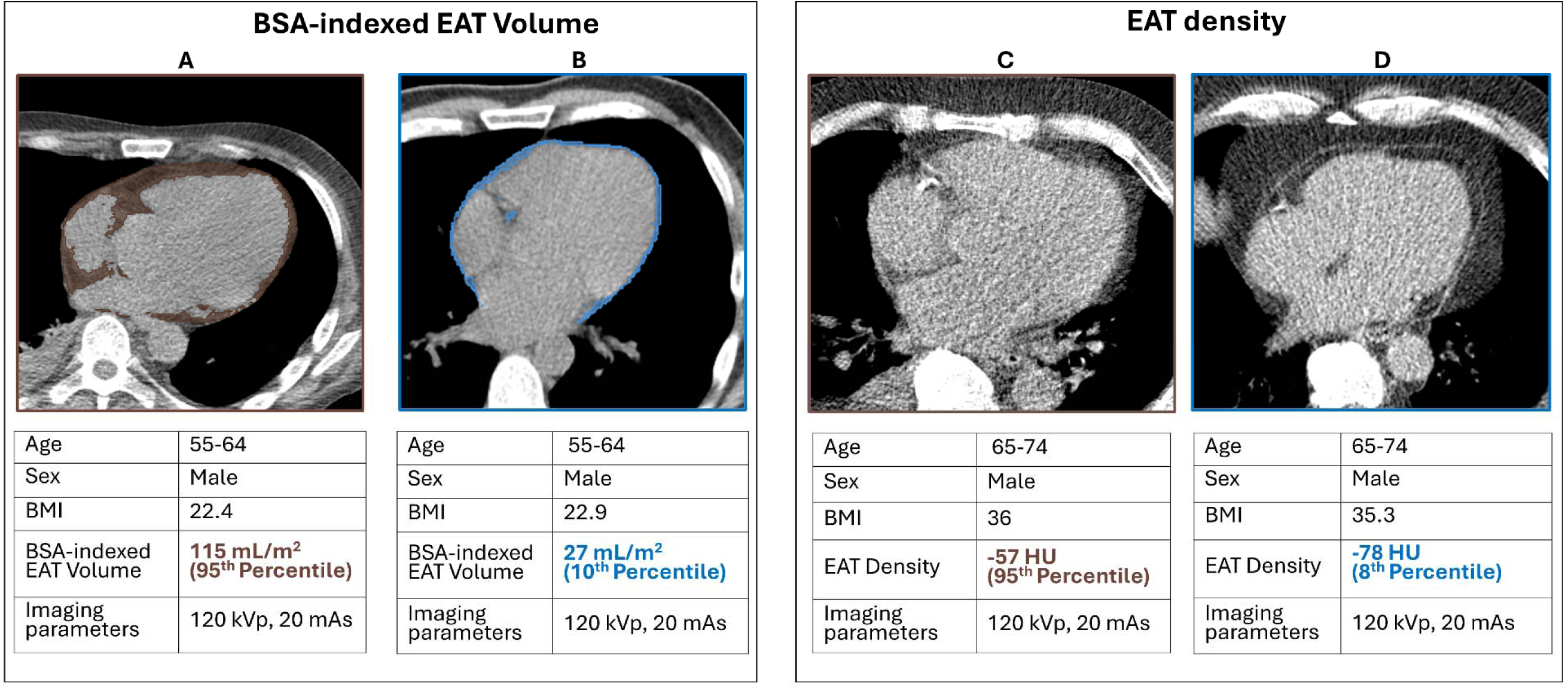
Case examples of patients within upper and lower percentiles of EAT density and BSA-indexed EAT volume. Panel A demonstrates a male in his 60s with a BMI of 22.4. His BSA-indexed EAT volume was within the 95th percentile for his age and sex group. He died during follow-up. Panel B demonstrates a male in his 60s with a BMI of 22.9. His BSA-indexed EAT volume was within the 10th percentile for his age and sex group. He remained event-free during follow-up. Panel C demonstrates a male in his 70s with a BMI of 36. His EAT density was within the 95th percentile for his age and sex group. He died during follow-up. Panel D demonstrates a male in his 70s with a BMI of 35.3. His EAT density was within the 8th percentile for his age and sex group. He remained event-free during follow-up

## DISCUSSION

Although AI now enables fully automated EAT quantification in routine clinical practice, clinicians lack an accepted framework to determine whether an individual patient’s EAT measurement is normal or abnormal. Unlike CAC, where percentile nomograms are routinely used for interpretation, no comparable reference standards exist for EAT. This gap is addressed in this study by developing and externally validating demographic-specific percentile nomograms in more than 42,000 patients. We also demonstrate that higher demographic-specific percentiles identify patients at progressively greater risk of death or myocardial infraction even after adjusting for myocardial perfusion parameters, clinical history, imaging heterogeneity, and CAC score. Importantly, utilizing percentile thresholds provides a meaningful mechanism for communicating EAT results; enabling clinicians to translate the insights regarding inflammation and cardiometabolic risk into clinical practice

We developed percentile representation to characterize the continuous relationship between EAT measurements and cardiovascular risk. In contrast, previous studies have primarily relied on binary cutoffs to define abnormal EAT values for risk prediction, providing limited insight into how cardiovascular risk changes across the continuous spectrum of EAT measurements [12, 15, 18]. The percentile approach has been used extensively for CAC scoring results and is currently being applied to coronary plaque volumes [21, 22]. McClelland et al. demonstrated substantial differences in CAC distributions by constructing sex-, age-, and race-specific CAC percentile nomograms in asymptomatic individuals. Their work showed that fixed global thresholds fail to account for important demographic variation, supporting the use of percentile-based interpretation rather than universal cutoffs. Prior studies have demonstrated that EAT also varies significantly by sex and race. In a study examining the changes in EAT measurements of 24,008 patients from the National Lung Screening Trial, Brendel et al. demonstrated that overall, women tended to have lower EAT volume and higher EAT density than men and that increased EAT median density and volume were associated with greater risk for all-cause mortality[18]. These population-specific differences underscore the value of percentile-based thresholds, which account for expected variation in EAT measures across demographic groups and allow for more meaningful risk stratification. Our findings are consistent with these prior observations, women across each sex-and age specific quartiles exhibit higher median EAT density than men, but lower BSA-indexed EAT volume than men. We also demonstrate that across both sexes, EAT median density decreases with age while EAT BSA-indexed volume increases with age. Additionally, prior studies reported ethnic differences in EAT. Adams et al. demonstrated significant differences in the epicardial fat volume between south Asians, southeast or east Asians, and Caucasian participants. South Asian patients had the highest median epicardial volume, while Caucasian patients had the lowest [25]. In our study, we observed significant differences in EAT density and BSA-indexed EAT volume between White and Black patients. When stratified by age, Black patients had higher median EAT density and lower EAT BSA-indexed EAT volume. However, race-, sex-, and age-stratified analyses demonstrated more complex EAT distributions at higher percentiles, suggesting potential interaction effects between demographic variables. Overall, these results suggest that EAT density and volume percentiles could provide both a mechanism to evaluate risk beyond binary classification as well as an intuitive way to communicate results with referring physicians and patients, who may be less familiar with EAT measurements.

In this large multicenter study, EAT density was a stronger predictor of cardiovascular risk then EAT volume across sex-, age-, and race-specific thresholds, with patients in the >95^th^ percentile exhibiting a 70% higher risk of adverse cardiovascular events compared with those below the 50th percentile. Unlike EAT volume, which primarily reflects abnormal metabolic states leading to ectopic fat deposition, EAT density may better capture its current biological state, including inflammatory and fibrotic remodeling. Prior studies have demonstrated that increased CT attenuation of pericoronary adipose tissue is associated with coronary inflammation and risk of plaque rupture, supporting the concept that CT-derived adipose tissue attenuation reflects tissue composition and biological activity rather than fat quantity alone [26, 27]. While pericoronary adipose tissue represents a regional component of EAT, these observations still provide biological plausibility for the stronger prognostic performance of EAT density observed in the present study.[8, 9, 28–30]. Additionally, the clear pathophysiologic link to coronary inflammation suggests it may have an important role as a surrogate biomarker in anti-inflammatory therapy trials [31, 32].

Our work, and in particular BMI subgroup analyses, also provides insights into the biological differences between EAT volume and EAT density. One possible explanation for the stronger prognostic value of EAT density is that the processes driving increased attenuation are different from those leading to ectopic fat deposition [33–35]. Increased EAT volume was common among obese individuals, where cardiometabolic risk factors are prevalent. As a result, EAT volume may provide limited discrimination in patients with a higher BMI. Conversely, among individuals with normal BMI, increased EAT volume may identify patients with increased cardiometabolic risk (leading to ectopic fat deposition) that would otherwise be underestimated using conventional anthropometric measures alone. In contrast, EAT density may capture inflammatory and fibrotic remodeling of epicardial fat, as outlined above, thereby providing additional prognostic information beyond generalized adiposity. Together, these findings suggest that EAT volume and EAT density provide complementary information that could help guide clinical management decisions.

While our findings highlight the potential of EAT quantification to identify patients at increased cardiovascular risk, these measurements should be interpreted within the broader clinical context to guide patient management. The significant differences in distribution across age, sex, and race highlight the need to move beyond single thresholds for EAT quantification. Percentiles provide an intuitive method for translating results into a patient-specific metric which also provides insights into the relative magnitude of elevation. Such simplified tools for clinical translation are needed in the context of an increasing number of biomarkers that could be used to tailor individual treatments.

Originally, EAT quantification has been proposed from gated cardiac CT scans, which synchronize images with the cardiac cycle and reduce susceptibility to motion artifacts seen in ungated CT imaging [36]. However, recent studies showed that EAT volume and density can be automatically quantified from ungated CT attenuation correction scans obtained during SPECT and PET [12]. Because low-dose ungated CT attenuation correction scans are routinely acquired during nuclear cardiology imaging, EAT may serve as an opportunistic imaging biomarker for cardiovascular risk stratification without requiring additional imaging.

### Limitations

There are a few important limitations present in our study. Firstly, it is a retrospective observational study, therefore we cannot directly conclude that high EAT density or volume causes adverse cardiovascular outcomes. Our percentile thresholds were also calculated based on a population of symptomatic patients referred to cardiac functional imaging (PET or SPECT), and as such cannot be readily generalized to asymptomatic patients or be used as a biomarker for death or MI risk in the general population. Lastly, all 42,842 AI-derived EAT measurements were not manually reviewed by an expert reader. However, given the large size of the multicenter cohort, manual evaluation of every CTAC scan would require a significant amount of time and resources.

### Conclusions

We demonstrate that EAT density and volume vary systematically according to age, sex, and race, with EAT density emerging as the stronger predictor of adverse cardiovascular events. By introducing externally validated demographic-specific percentile nomograms, this study provides a practical reference framework for translating automated EAT measurements into clinically meaningful risk estimates. This approach may facilitate precision risk assessment by identifying high-risk individuals whose risk is not fully captured by conventional clinical risk parameters.

## FUNDING

This research was supported in part by grant R35HL161195 from the National Heart, Lung, and Blood Institute/National Institutes of Health (NHLBI/NIH) and R01EB034586 from the National Institute of Biomedical Imaging and Bioengineering (PI: Piotr Slomka). The content is solely the responsibility of the authors and does not necessarily represent the official views of the National Institutes of Health.

## DISCLOSURE OF INTEREST

RM received consulting fees from Alnylam and Bayer and research support from Alberta Innovates. DB and PS participated in software royalties for QPS software at Cedars-Sinai Medical Center. DB, DD, and PS reported equity in APQ Health Inc. DB received research grant support from The Dr. Miriam and Sheldon G. Adelson Medical Research Foundation and consulting fees from GE Healthcare. PS received research grant support from Siemens Medical Systems, and consulting fees from Synektik SA and Novo Nordisk. PC reported consulting for Clario. MDC reported consulting fees from MedTrace, Valo Health, GE, Bitterroot Bio, and IBA, investigator-initiated research support from Amgen, and institutional research grant support from Sun Pharma, Xylocor, Alnylam, and Intellia. AJE reports consulting for Artrya, authorship fees from Wolters Kluwer Healthcare—UpToDate, and serving on scientific advisory boards for Axcellant and Canon Medical Systems USA; his institution has grants/grants pending from Alexion, Attralus, BridgeBio, Canon Medical Systems USA, GE HealthCare, Intellia Therapeutics, International Atomic Energy Agency, Ionis Pharmaceuticals, National Institutes of Health, Pfizer, and Shockwave Medical. RRSP serves as a consultant for GE HealthCare. MA-M received research support from Siemens and GE Healthcare and is a consultant to Jubilant, Medtrace, GE Healthcare, and Pfizer. LS received grant support/consulting honorarium from Amgen and Philips and served as 3 site PI for V-INITIATE and Ocean(a) trials. VTL has received research grant support from J&J/Janssen, has received honorarium from the American College of Cardiology for Editor-inChief role at Cardiosmart, and has served on advisory boards for Amgen, Amarin, Bayer, Boehringer Ingelheim, Esperion, Idorsia, iRhythm, Merck, Novartis, Novonordisk, and Pfizer. RBP reports receiving honoraria as a Section Editor for UpToDate and stipends for serving as an Associate Editor for Circulation: Cardiovascular Imaging and JACC: Advances The remaining authors declare no conflict of interest

## DATA AVAILABLITY

To the extent allowed by data sharing agreements and IRB protocols, the deidentified data and data analysis code from this manuscript will be shared upon written request.

## Data Availability

https://staging.dqkgxywpqmaju.amplifyapp.com/

## Notes

### Author Declarations

The study was approved by the institutional review boards (IRB) at each participating center, and the overall study was approved by the Cedars-Sinai Medical Center institutional review board. The study compiled with the Declaration of Helsinki. At each participating center, imaging and clinical data were linked locally under site-specific IRB approval before assignment of a unique study ID and deidentification. Centers obtained either written informed consent or a waiver of consent for retrospective data, according to site-specific protocols. Only deidentified data were transferred to the core laboratory; all protected health information, including identifiers within image files, was removed using HIPAA-compliant software prior to transfer.

