## Supplemental Material for "Clinical Reference Percentiles for AI-derived Epicardial Adipose Tissue: A Multicenter Study"

**Supplemental Tables**

Supplemental Table 1	2

Supplemental Table 2	3

Supplemental Table 3	5

Supplemental Table 4	6

Supplemental Table 5	8

Supplemental Table 6	9

**Supplemental Figures**

Supplemental Figure 1	10

Supplemental Figure 2	11

Supplemental Figure 3	12

Supplemental Figure 4	13

Supplemental Figure 5	14

Supplemental Figure 6	15

Supplemental Figure 7	17

Supplemental Figure 8	18

Supplemental Figure 9	19

Supplemental Figure 10	21

Supplemental Figure 11	23

Supplemental Figure 12	25

Supplemental Table 1

|  | **Site** | **N** | **Imaging Modality** |
| --- | --- | --- | --- |
| **Derivation N = 15,082** | Brigham and Women's Hospital | 7,058 | PET |
|  | Ottawa | 5,115 | SPECT & PET |
|  | Houston Methodist | 1,892 | PET |
|  | West Los Angeles Veterans Affairs | 1017 | PET |
| **Validation**  **N= 27,760** | University of Calgary | 2,912 | SPECT |
|  | Cedars- Sinai Medical Center | 5,945 | PET |
|  | Columbia | 3,787 | PET & SPECT |
|  | Intermountain Healthcare | 5,685 | PET |
|  | Kansas University Medical Center | 590 | PET |
|  | Mayo Clinic | 1,991 | PET |
|  | Mexico City | 364 | PET |
|  | Montefiore Medical Center | 1,015 | PET |
|  | University of Naples | 456 | PET |
|  | Yale | 4,266 | SPECT |
|  | Zurich | 749 | PET |

Supplemental Table 1. Sites and imaging modalities of derivation and validation cohort. Abbreviations: PET – positron emission tomography, SPECT – single-photon emission computed tomography

Supplemental Table 2

| **Site** | **Modality** | **Scanner** | **Slice Thickness [mm]** | **Tube Current [mA]** | **Tube Voltage [kVp]** | **Breath hold** |
| --- | --- | --- | --- | --- | --- | --- |
| Brigham and Women's Hospital | PET | GE Discovery MI  GE Discovery RX  GE Discovery STE | 2.5-5 | 10-26 | 120-140 | Shallow |
| Calgary | SPECT | GE Discovery 570c | 5 | 16 | 120 | Free |
| Cedars-Sinai | PET | Siemens Biograph 64 TruePoint  Siemens Biograph 128 Vision Edge  GE Discovery 710 | 3 | 11-13 | 100 | Shallow |
| Columbia University Irving Medical Center | PET | Siemens Biograph 64 mCT Flow | 3 | 30 | 120 | Free |
| Columbia University Irving Medical Center | SPECT | Phillips Precedence 16P | 5 | 16 | 120 | Free |
| Houston Methodist Academic Institute | PET | Siemens Biograph 600 Vision Edge | 3 | 20-50 | 100-120 | Free |
| Intermountain Medical Center | PET | Intermountain Medical Center | 2 | 15-38 | 120-130 | Free |
| University of Kansas Medical Center | PET | GE Discovery MI | 3.75 | 14-75 | 120 | Free |
| Montefiore Medical Center | PET | Philips Gemini TF TOF 16  Philips Gemini TF TOF 64 | 3 | 110-185 | 120 |  |
| Mayo Clinic | PET | GE Discovery 710 | 3.75 | 17-77 | 120 | Shallow |
| National Autonomous University of Mexico | PET | Siemens Biograph 64 TruePoint  Siemens Biograph Vision 600 | 3 | 33-580 | 120 | Breathe Hold |
| University of Naples Federico II | PET | Philips Ingenuity TF  GE Discovery MI | 3 | 35-100 | 120-140 | Free |
| Ottawa Heart Institute | SPECT | Siemens Symbia Intevo 16 | 5 | 20 | 120 | Free |
| West Los Angeles Veterans Affair Medical Center | PET | Siemens Biograph 64 mCT  Siemens Biograph 64 Vision 600 | 3 | 70-200 | 120 | Free |
| Yale | SPECT | GE Discovery 570c | 2.5 | 60-150 | 120 | Breathe Hold |
| University Hospital Zurich | PET | GE Discovery STE  GE Discovery RX  GE Discovery LS  GE Discovery HR | 2.5-5 | 29-350 | 100-140 | Free |

Supplemental Table 2: Acquisition parameters for computed tomography scans used in the study. Abbreviations: PET – positron emission tomography, SPECT – single-photon emission computed tomography

Supplemental Table 3

| **Variable** | **Overall**  **N = 42,842** | **Derivation**  **N = 15,082** | **Validation**  **N = 27,760** |
| --- | --- | --- | --- |
| Race | 8,066 (19%) | 2,539 (17%) | 5,527 (20%) |
| Stress ejection fraction | 189 (0.44%) | 14 (0.1%) | 175 (0.6%) |

Supplemental Table 3. Data missingness. Only variables for which data was missing are presented.

Supplemental Table 4

| **Characteristic** | **Overall**  N = 42,842 | **Female**  N = 17,953 | **Male**  N = 24,889 | **P value** |
| --- | --- | --- | --- | --- |
| Age | 67.0 (58.0, 75.0) | 67.0 (58.0, 75.0) | 67.0 (58.0, 75.0) | <0.001 |
| Race |  |  |  | <0.001 |
| *American Indian or Alaska Native* | 157 (0.4%) | 76 (0.4%) | 81 (0.3%) |  |
| *Asian* | 1,147 (2.7%) | 441 (2.5%) | 706 (2.8%) |  |
| *Black or African American* | 4,640 (11%) | 2,445 (14%) | 2,195 (8.8%) |  |
| *Native Hawaiian or Other Pacific Islander* | 199 (0.5%) | 92 (0.5%) | 107 (0.4%) |  |
| *Unknown* | 8,066 (19%) | 3,778 (21%) | 4,288 (17%) |  |
| *White* | 28,633 (67%) | 11,121 (62%) | 17,512 (70%) |  |
| BMI | 29.4 (25.5, 34.6) | 30.1 (25.2, 35.9) | 29.0 (25.6, 33.7) | <0.001 |
| BSA (m²) | 2.0 (1.8, 2.2) | 1.8 (1.7, 2.0) | 2.1 (1.9, 2.2) | <0.001 |
| Hypertension | 31,747 (74%) | 13,017 (73%) | 18,730 (75%) | <0.001 |
| Dyslipidemia | 29,022 (68%) | 11,395 (63%) | 17,627 (71%) | <0.001 |
| Diabetes Mellitus | 14,499 (34%) | 5,770 (32%) | 8,729 (35%) | <0.001 |
| Family History of CAD | 11,940 (28%) | 5,418 (30%) | 6,522 (26%) | <0.001 |
| Past Myocardial Infarction | 7,131 (17%) | 2,173 (12%) | 4,958 (20%) | <0.001 |
| Smoking | 10,027 (23%) | 3,495 (19%) | 6,532 (26%) | <0.001 |
| CAC score | 139.6 (0.0, 977.9) | 24.4 (0.0, 348.4) | 357.4 (17.2, 1,514.7) | <0.001 |
| Left ventricle volume (mL) | 122.5 (100.1, 151.2) | 101 (86.5, 119.7) | 139.5 (118.3, 168.0) | <0.001 |
| EAT volume (mL) | 112.6 (74.6, 161.2) | 95.4 (64, 137) | 126.2 (85.3, 176.9) | <0.001 |
| EAT indexed BSA (mL/m²) | 57.4 (39.3, 79.7) | 52.8 (35.8, 74.3) | 61.1 (42.3, 83.1) | <0.001 |
| EAT % heart size | 19.5 (13.1, 27.7) | 20.2 (13.4, 29.0) | 19.0 (12.9, 26.8) |  |
| EAT density (HU) | -67 (-71, -62) | -66 (-71, -61) | -67 (-72, -62) | <0.001 |

Supplemental Table 4. Patient demographics stratified by Sex. Categorical variables are shown as n (%) and were compared with Pearson’s Chi-squared test, while continue variables are shown as median, Interquartile range (IQR) and were compared with Wilcoxon rank sum test. Abbreviations: BMI – body mass index, BSA – body surface area, CAD – coronary artery disease, EAT – epicardial adipose tissue, HU – Hounsfield unit, CAC – coronary calcium artery score

Supplemental Table 5

| **Sex** | **Age** | **10^th^ Percentile** | **25^th^ Percentile** | **50^th^ Percentile** | **75^th^ Percentile** | **90^th^ Percentile** | **95^th^ Percentile** |
| --- | --- | --- | --- | --- | --- | --- | --- |
| Male | 40 | 21.1 (20.2, 22) | 31.2 (30, 32.2) | 44.4 (42.8, 45.9) | 60.8 (58.7, 62.5) | 79.2 (76.5, 81.5) | 93 (89.5, 96) |
|  | 50 | 23.5 (22.7, 24.3) | 34.6 (33.7, 35.4) | 48.3 (48, 50.5) | 67.4 (65.9, 68.7) | 87.8 (85.6, 89.7) | 103.1 (100.2, 105.6) |
|  | 60 | 26.1 (25.4, 26.9) | 38.5 (37.6, 39) | 54.7 (53.8, 55.6) | 74.8 (73.6, 75.7) | 97.4 (95.5, 98.9) | 114.3 (111.8, 116.5) |
|  | 70 | 29.1 (28.4, 29.8) | 42.7 (41.9, 43.4) | 60.7 (60, 61.6) | 83 (81.8, 84) | 108 (106, 109.6) | 126.7 (124.2, 129) |
|  | 80 | 32.3 (31.4, 33.2) | 47.4 (46.3, 48.4) | 67.3 (66.3, 68.7) | 92 (90.2, 93.7) | 119.7 (116.9, 122) | 140.5 (137.2, 143.5) |
| Female | 40 | 16.5 (15.6, 17.2) | 24.5 (23.6, 25.5) | 36.1 (34.7, 37.6) | 50.5 (48.6, 52.6) | 68.2 (65.3, 71.2) | 81.1 (77.7, 84.5) |
|  | 50 | 18.9 (18.1, 19.5) | 28.1 (27.2, 28.9) | 41.2 (40, 42.4) | 57.6 (56, 59.4) | 77.8 (75.2, 80.4) | 92.4 (89.3, 95.6) |
|  | 60 | 21.6 (20.9, 22.3) | 32.1 (31.3, 32.7) | 47.1 (46.2, 48) | 65.6 (64.4, 67.1) | 88.6 (86.4, 91.1) | 105.3 (102.2, 108.2) |
|  | 70 | 24.7 (24, 25.5) | 36.6 (35.8, 37.2) | 53.7 (52.8, 54.6) | 74.8 (73.5, 76.2) | 101 (98.8, 103.5) | 119.9 (116.6, 123.1) |
|  | 80 | 28.3 (27.3, 29.2) | 41.8 (40.7, 42.8) | 61.2 (60, 62.5) | 85.3 (83.5, 87.2) | 115.1 (112.2, 118.4) | 136.6 (132.9, 140.6) |

Supplemental Table 5. Sex-and age specific BSA-indexed EAT Volume Percentiles with 95% confidence intervals

Supplemental Table 6

| **Sex** | **Age** | **10^th^ Percentile** | **25^th^ Percentile** | **50^th^ Percentile** | **75^th^ Percentile** | **90^th^ Percentile** | **95^th^ Percentile** |
| --- | --- | --- | --- | --- | --- | --- | --- |
| Male | 40 | -73.1 (-73.6, -72.7) | -68.3 (-68.7, -68) | -64.1 (-64.4, -63.7) | -59.5 (-59.9, -59.2) | -55.9 (-56.2, -55.5) | -53.7 (-54.1, -53.3) |
|  | 50 | -74.2 (-74.6, -73.9) | -69.4 (-69.7, -69.2) | -65.2 (-65.4, -64.9) | -60.7 (-60.9, -60.4) | -57 (-57.3, -56.7) | -54.8 (-55.2, -54.5) |
|  | 60 | -75.3 (-75.7, -75) | -70.6 (-70.7, -70.4) | -66.3 (-66.5, -66.1) | -61.8 (-62, -61.6) | -58.1 (-58.3, -57.9) | -55.9 (-56.3, -55.7) |
|  | 70 | -76.4 (-76.8, -76.2) | -71.7 (-71.9, -71.5) | -67.4 (-67.6, -67.2) | -62.9 (-63.1, -62.7) | -59.2 (-59.4, -59) | -57.1 (-57.4, -56.8) |
|  | 80 | -77.6 (-78, -77.2) | -72.8 (-73.1, -72.5) | -68.6 (-68.8, -68.3) | -64 (-64.3, -63.8) | -60.3 (-60.6, -60.1) | -58.2 (-58.5, -57.9) |
| Female | 40 | -72.1 (-72.7, -71.6) | -67.1 (-67.5, -66.6) | -62.1 (-62.6, -61.7) | -57.5 (-57.9, -57.1) | -54.1 (-54.6, -53.7) | -52.2 (-52.6, -51.8) |
|  | 50 | -73.5 (-73.9, -73.1) | -68.5 (-68.7, -68.1) | -63.5 (-63.9, -63.2) | -58.9 (-59.2, -58.6) | -55.5 (-55.8, -55.2) | -53.6 (-53.9, -53.3) |
|  | 60 | -74.9 (-75.3, -74.5) | -69.9 (-70.0, -69.6) | -64.9 (-65.1, -64.6) | -60.2 (-60.5, -60.0) | -56.9 (-57.1, -56.7) | -55.0 (-55.3, -54.7) |
|  | 70 | -76.3 (-76.7, -75.9) | -71.2 (-71.4, -71.0) | -66.3 (-66.5, -66.0) | -61.6 (-61.8, -61.4) | -58.3 (-58.5, -58.0) | -56.3 (-56.7, -56.0 |
|  | 80 | -77.7 (-78.1, -77.2) | -72.6 (-72.9, -72.3) | -67.7 (-68.0, -67.3) | -63.0 (-63.3, -62.7) | -59.7 (-60.0, -59.3) | -57.7 (-58.1, -57.4) |

Supplemental Table 6. Sex- and -age specific EAT median Density percentiles with 95% confidence interval

**SUPPLEMENTAL FIGURES**

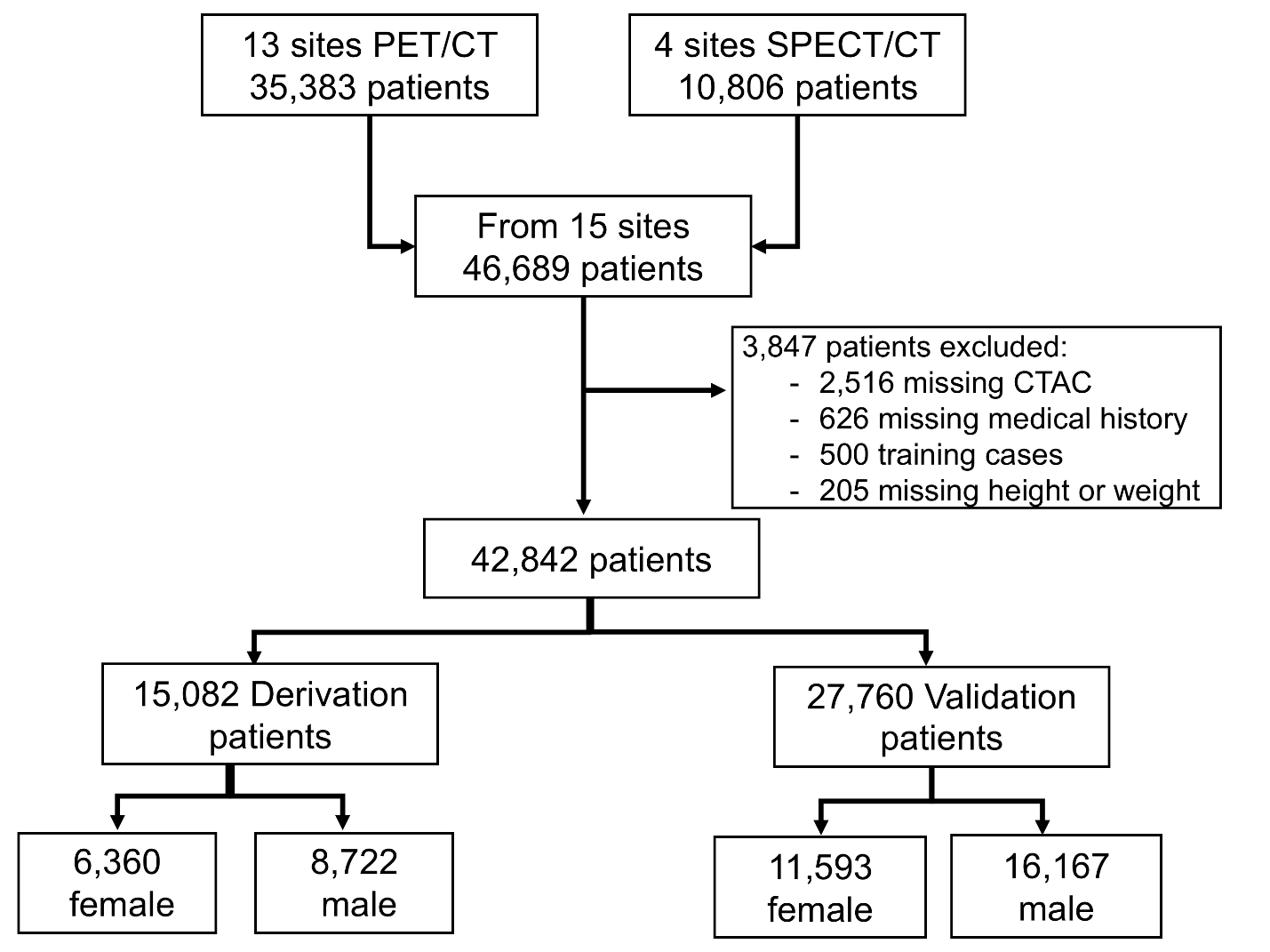
Supplemental Figure 1

Figure 1. Study design. 2 sites were included in both the REFINE SPECT and REFINE PET registry. 4 Sites comprising of both SPECT and PET modality were used for the derivation cohort. The remaining sites were used as the validation cohort. Abbreviations: PET/CT – positron emission tomography / computed tomography, SPECT – single-photon emission computed tomography, CTAC – computed tomography attenuation correction

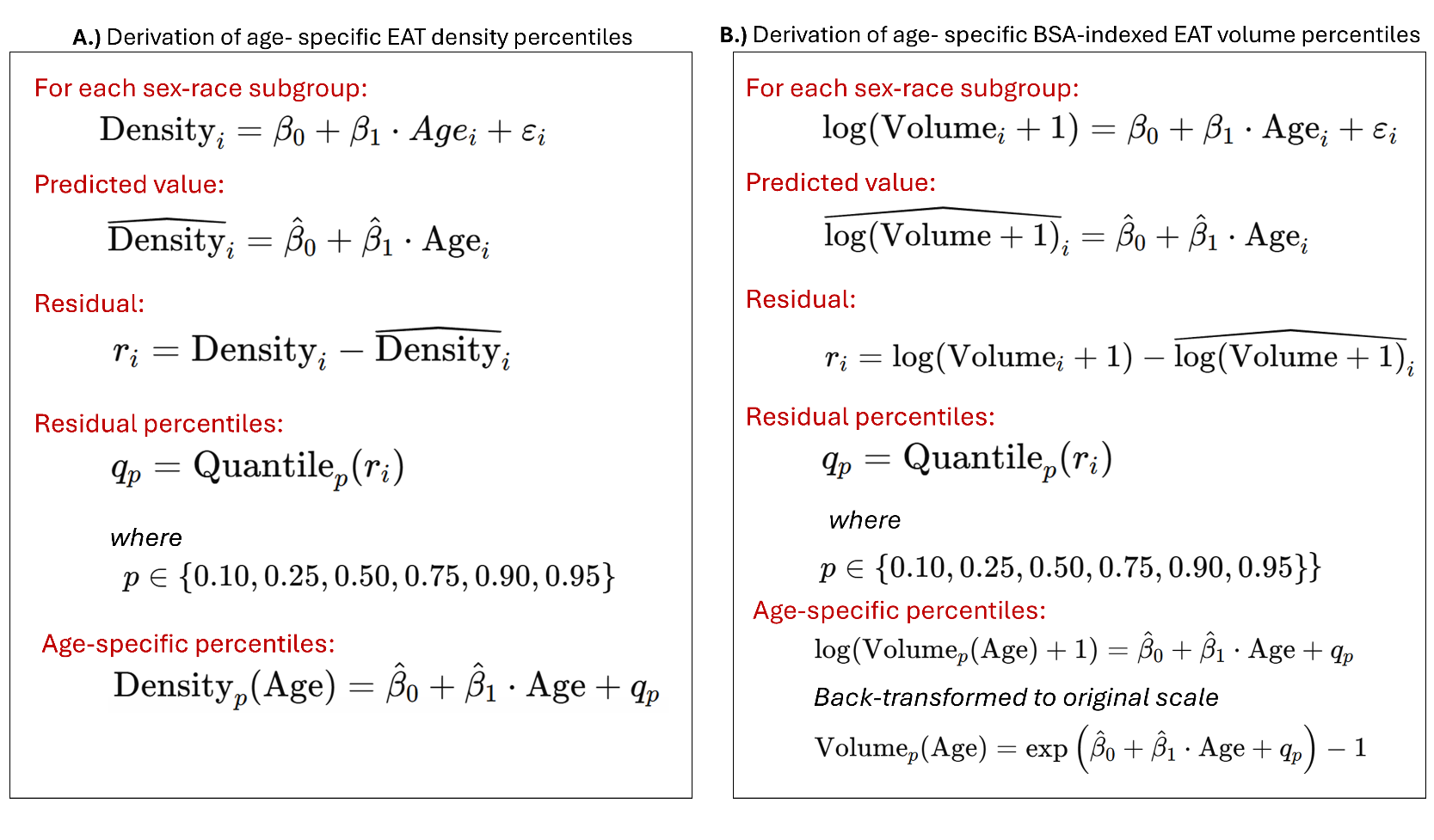
Supplemental Figure 2

Supplemental Figure 2. Panel A shows the derivation of age specific EAT density percentile curves. Panel B shows the derivation of BSA-indexed EAT volume percentile curves. For BSA-indexed EAT volume, percentile curves were derived on the log-transformed scale and back-transformed to the original scale.

Supplemental Figure 3

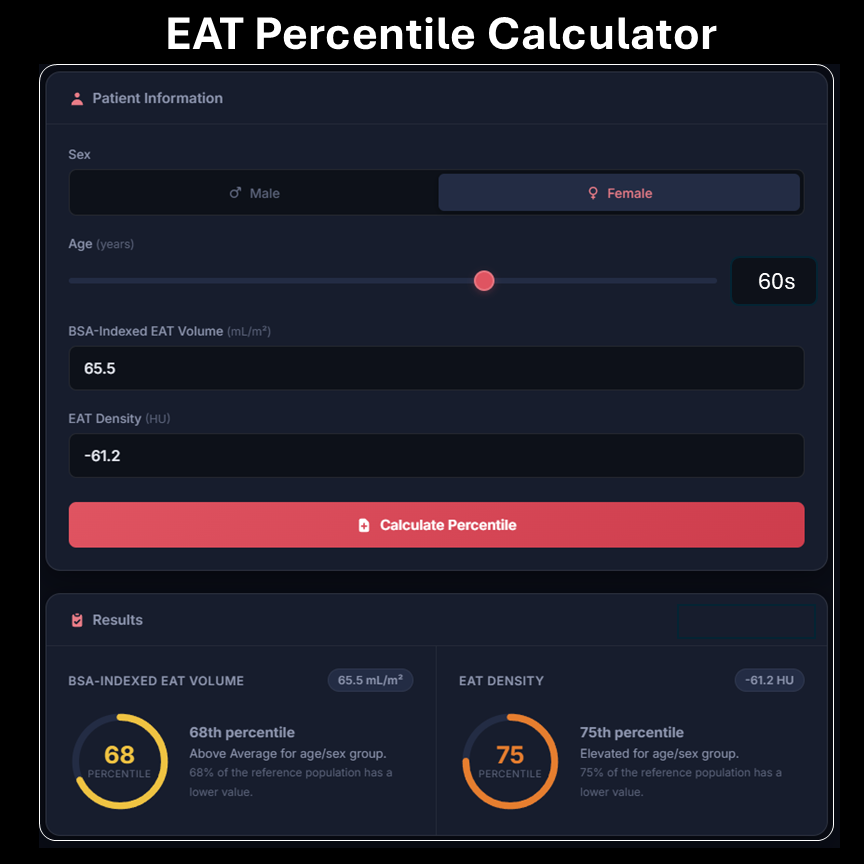

Supplemental Figure 3. This online EAT percentile calculator provides individualized sex-and age-specific percentiles for EAT density (Hounsfield Units) and BSA-indexed EAT volume (mL/m^2^). Percentiles were derived from a multicenter derivation cohort of 15,082 patients. In this example, a female in her 60s with a BSA-indexed EAT volume of 65.5 mL/m^2^ and an EAT density of -61.2 HU is in the 68^th^ and 75^th^ percentile, respectively, for her age and sex.

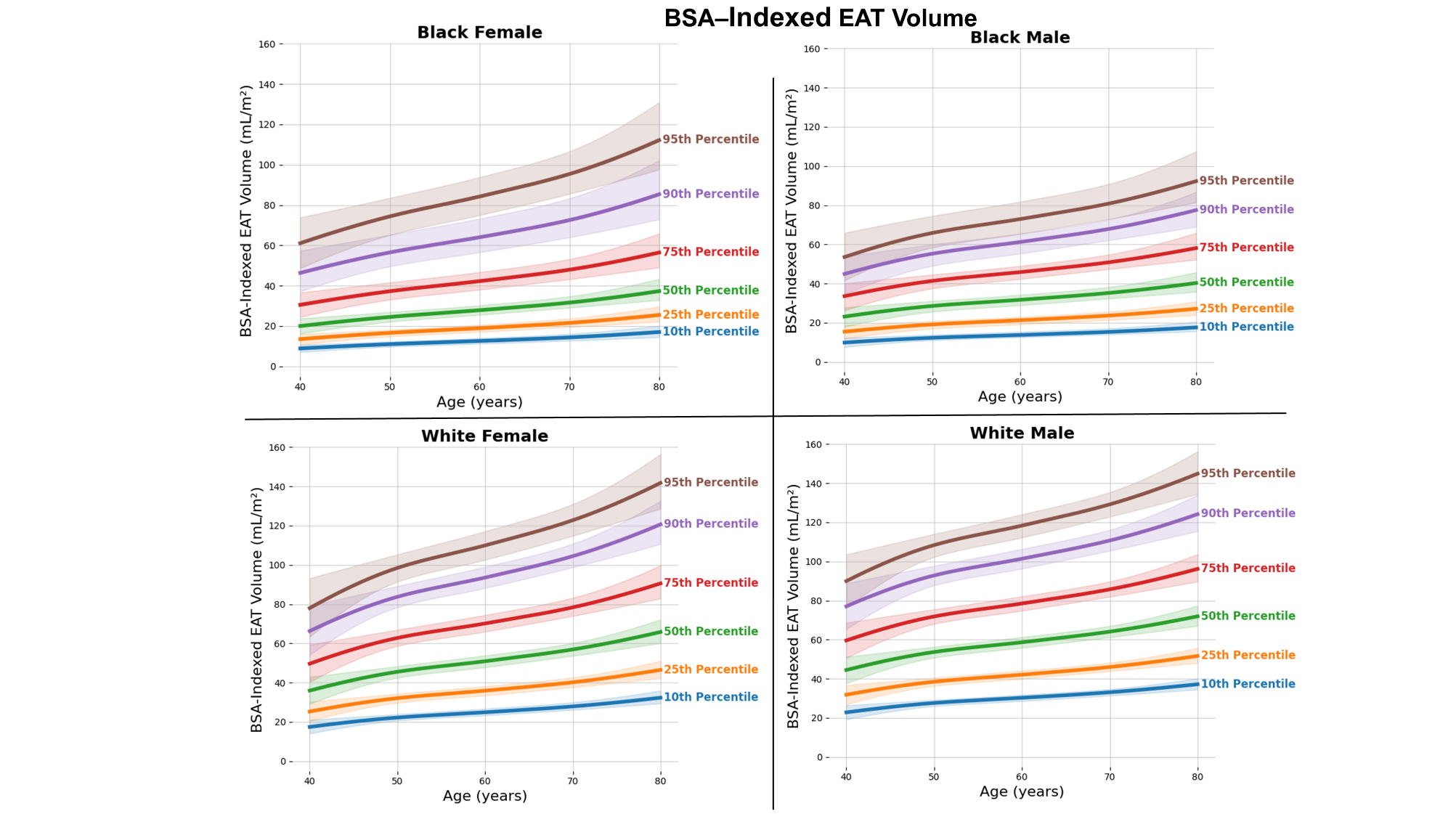
Supplemental Figure 4.

Supplemental Figure 4. Race-, sex-, and age-specific percentiles for BSA-indexed EAT volume with 95% confidence intervals.

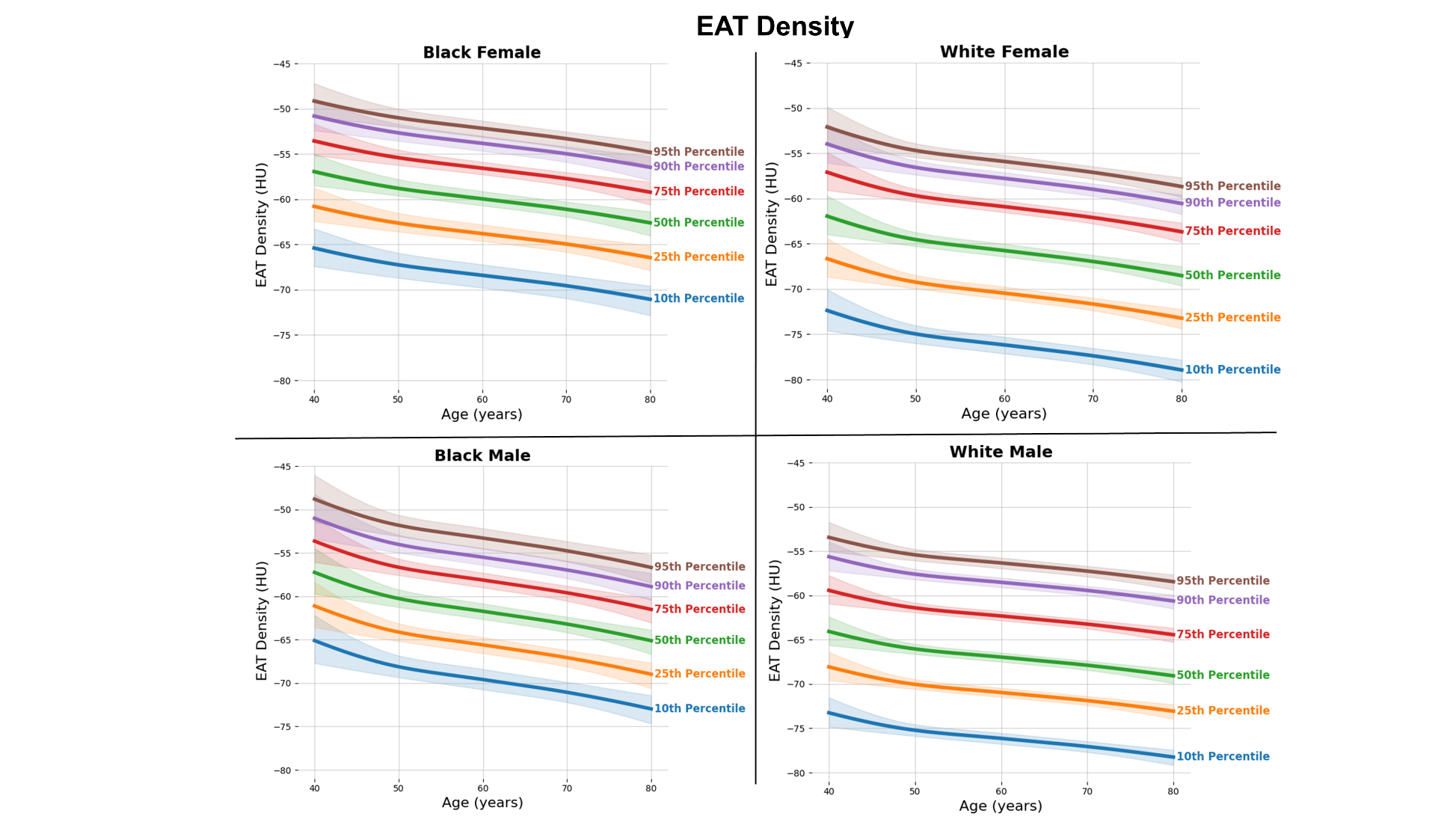
Supplemental Figure 5

Supplemental Figure 5. Race-, sex-, and age-specific percentiles for EAT density with 95% confidence intervals.

Supplemental Figure 6

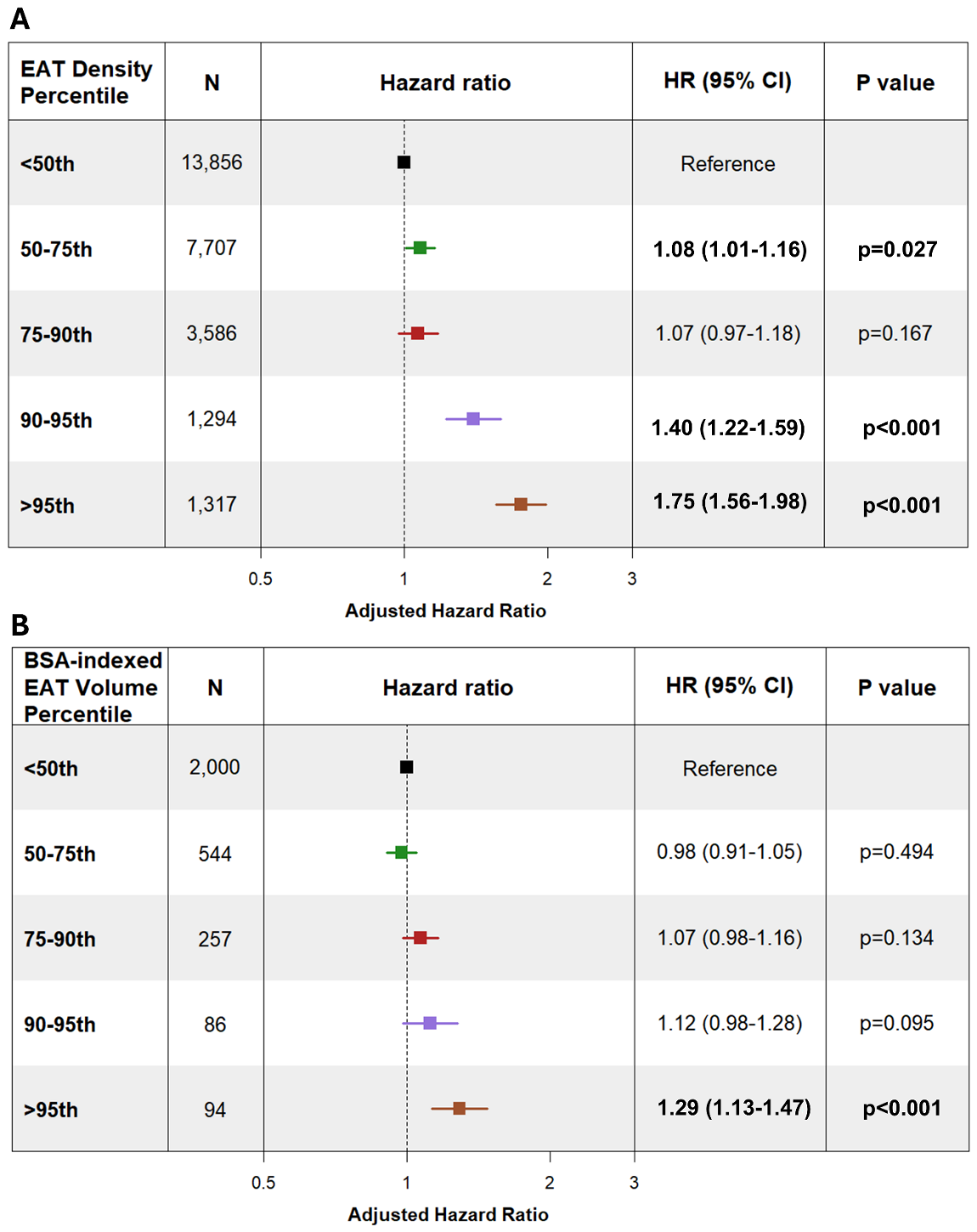

Supplemental Figure 6. Adjusted Cox proportional hazards model of sex- and age- stratified EAT percentile groups, with patients below the 50^th^ percentile serving as the reference group. Panel A displays adjusted hazard ratios across EAT density percentiles. Panel B displays adjusted hazard ratios across BSA-indexed EAT percentiles. Models were adjusted for tube current, tube voltage, site, race, dyslipidemia, diabetes mellitus, smoking, previous coronary artery disease, family history of coronary artery disease, body mass index, hypertension, past myocardial infarction, coronary artery calcium score, stress total perfusion deficient, and stress ejection fraction, along with imaging modality as shared frailty. N = 27,760.

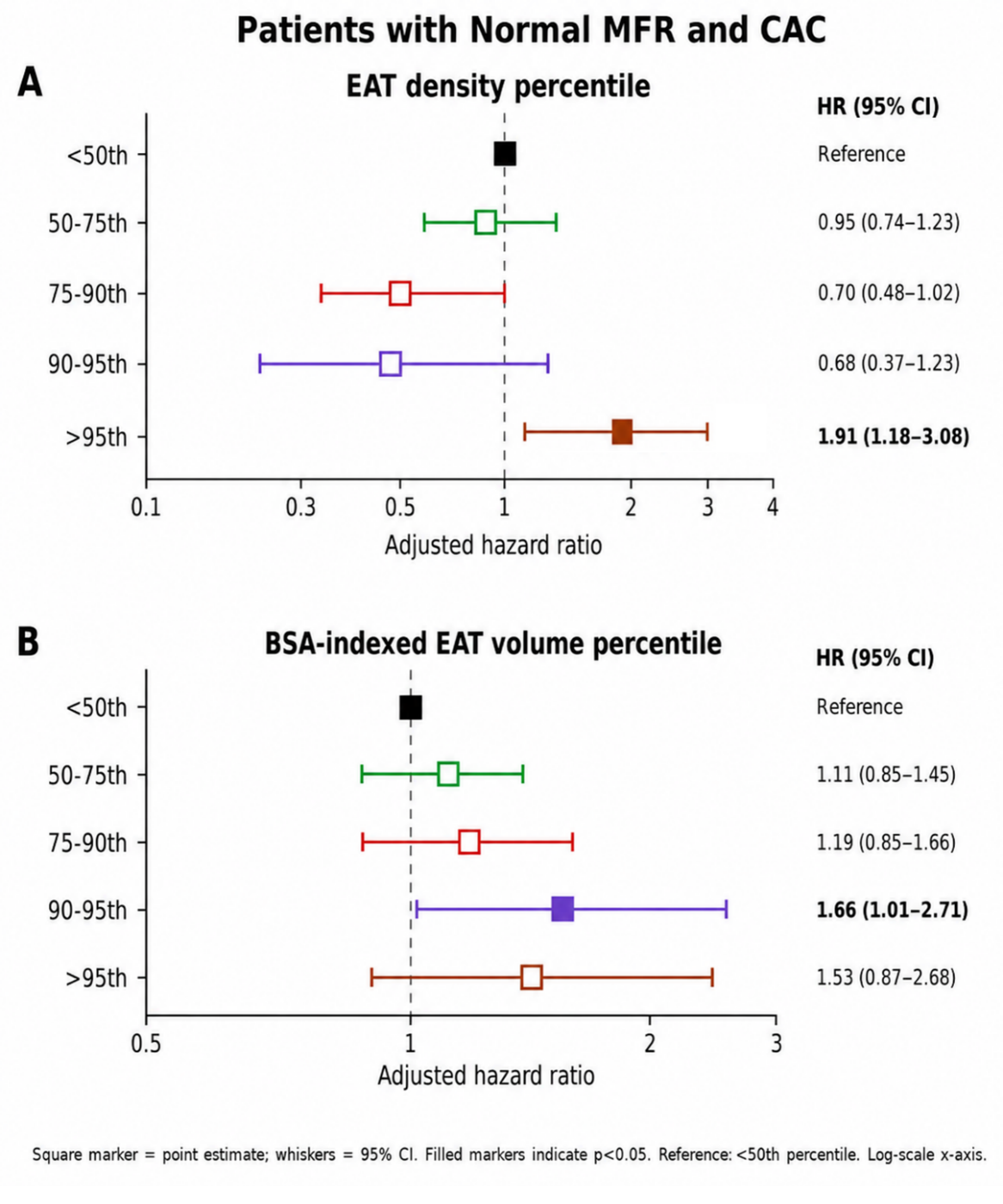
Supplemental Figure 7

Supplemental Figure 7. Adjusted Cox proportional hazards model of sex- and age- stratified EAT percentile groups in patients with myocardial flow reserve greater than 2 and a coronary artery calcium score of 0. Models were adjusted for dyslipidemia, diabetes mellitus, smoking, previous coronary artery disease, family history of coronary artery disease, hypertension, past myocardial infarction, and stress ejection fraction. Race and site were excluded from the multivariable model because sparse data within several categories resulted in unstable parameter estimates. N = 3,822

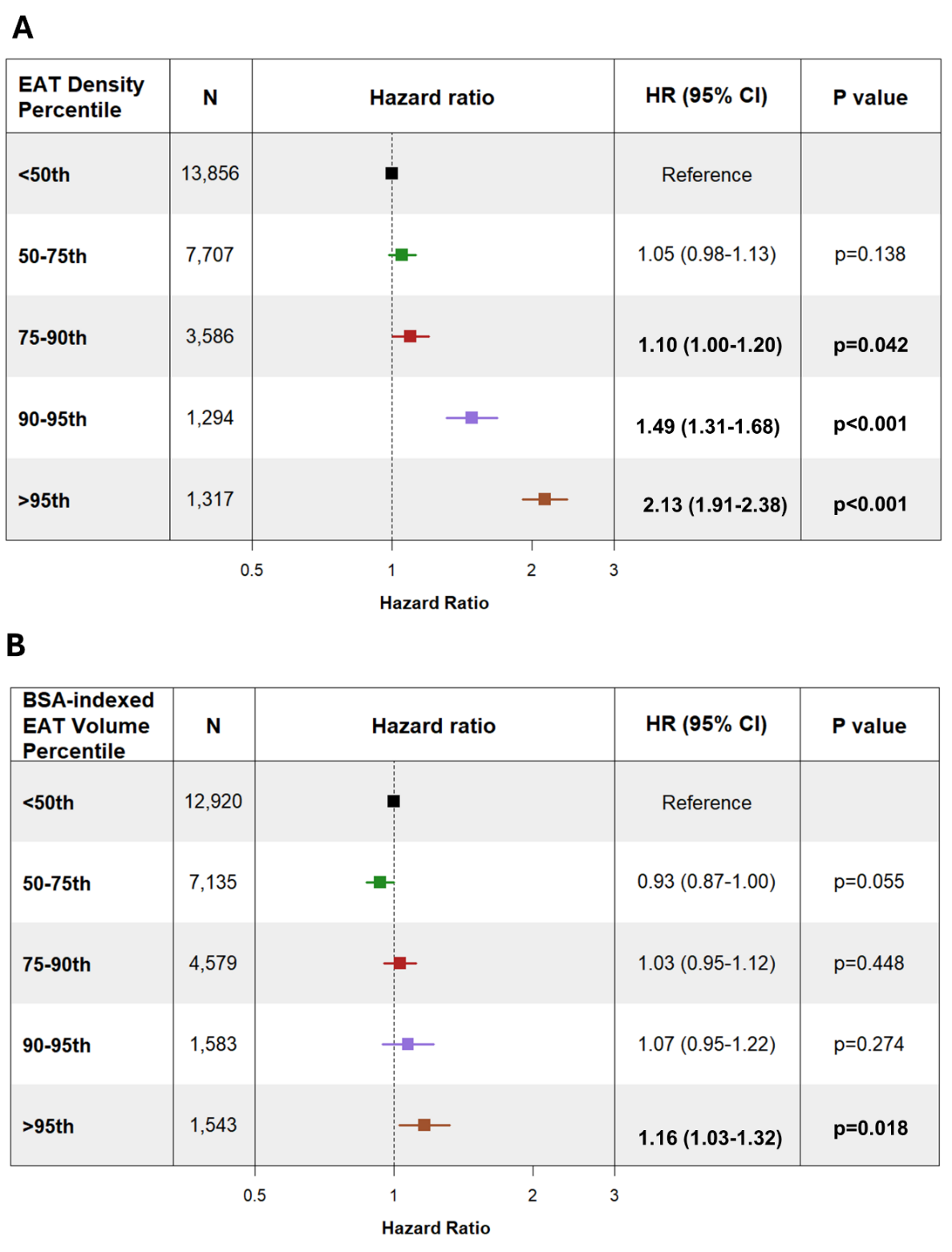
Supplemental Figure 8

Supplemental Figure 8. Unadjusted Cox proportional hazards model of sex- and age- stratified EAT percentile groups, with patients below the 50^th^ percentile serving as the reference group. Panel A displays unadjusted hazard ratios across EAT density percentiles. Panel B displays unadjusted hazard ratios across BSA-indexed EAT percentiles. N = 27,760.

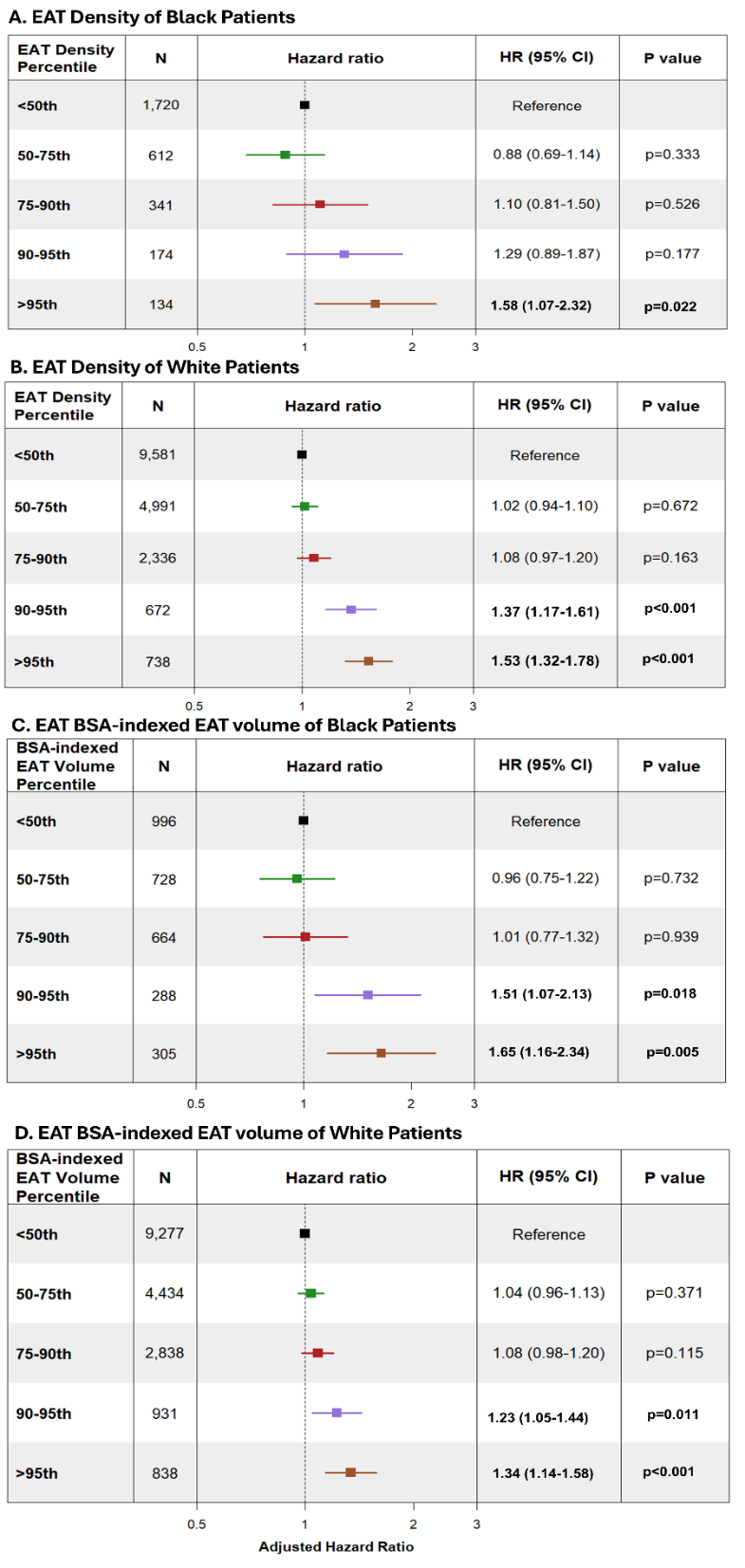
Supplemental Figure 9

Supplemental Figure 9. Adjusted Cox proportional hazards model of race-, sex-, and age- stratified EAT percentile groups, with patients under the 50^th^ percentile serving as the reference group. Panel A displays adjusted hazard ratios across EAT density percentiles in Black patients. Panel B displays adjusted hazard ratios across EAT density percentiles in White patients. Panel C displays adjusted hazard ratios across BSA-indexed EAT percentiles in Black patients. Panel D displays adjusted hazard ratios across BSA-indexed EAT percentiles in White patients. Models were adjusted for site, dyslipidemia, diabetes mellitus, smoking, previous coronary artery disease, family history of coronary artery disease, body mass index, hypertension, past myocardial infarction, coronary artery calcium score, stress total perfusion deficit, stress ejection fraction, along with imaging modality as shared frailty.

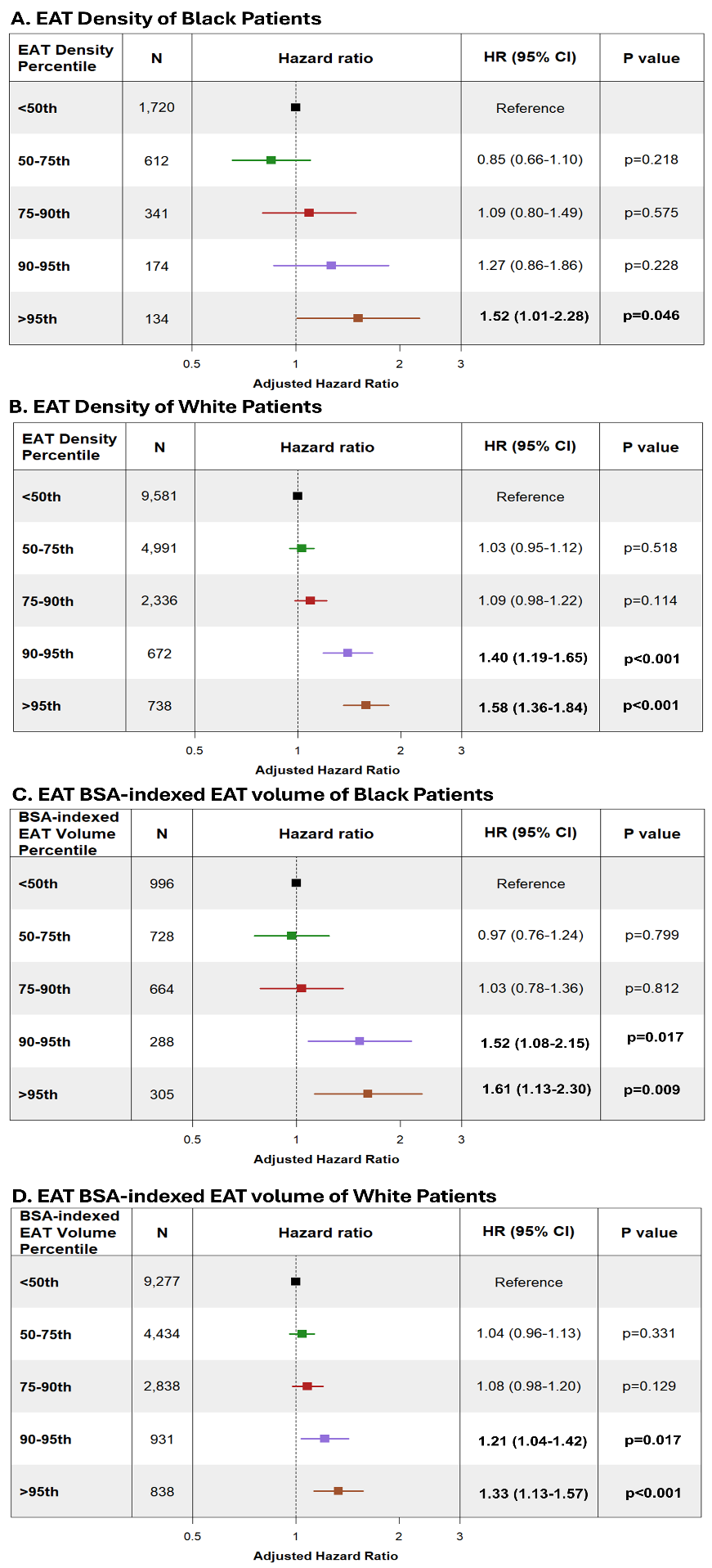
Supplemental Figure 10

Supplemental Figure 10. Adjusted Cox proportional hazards model of race-, sex-, and age- stratified EAT percentile groups, with patients under the 50^th^ percentile serving as the reference group. Models were adjusted for tube current, tube voltage, site, dyslipidemia, diabetes mellitus, smoking, previous coronary artery disease, family history of coronary artery disease, body mass index, hypertension, past myocardial infarction, coronary artery calcium score, stress total perfusion deficit, stress ejection fraction, along with imaging modality as shared frailty.

Supplemental Figure 11

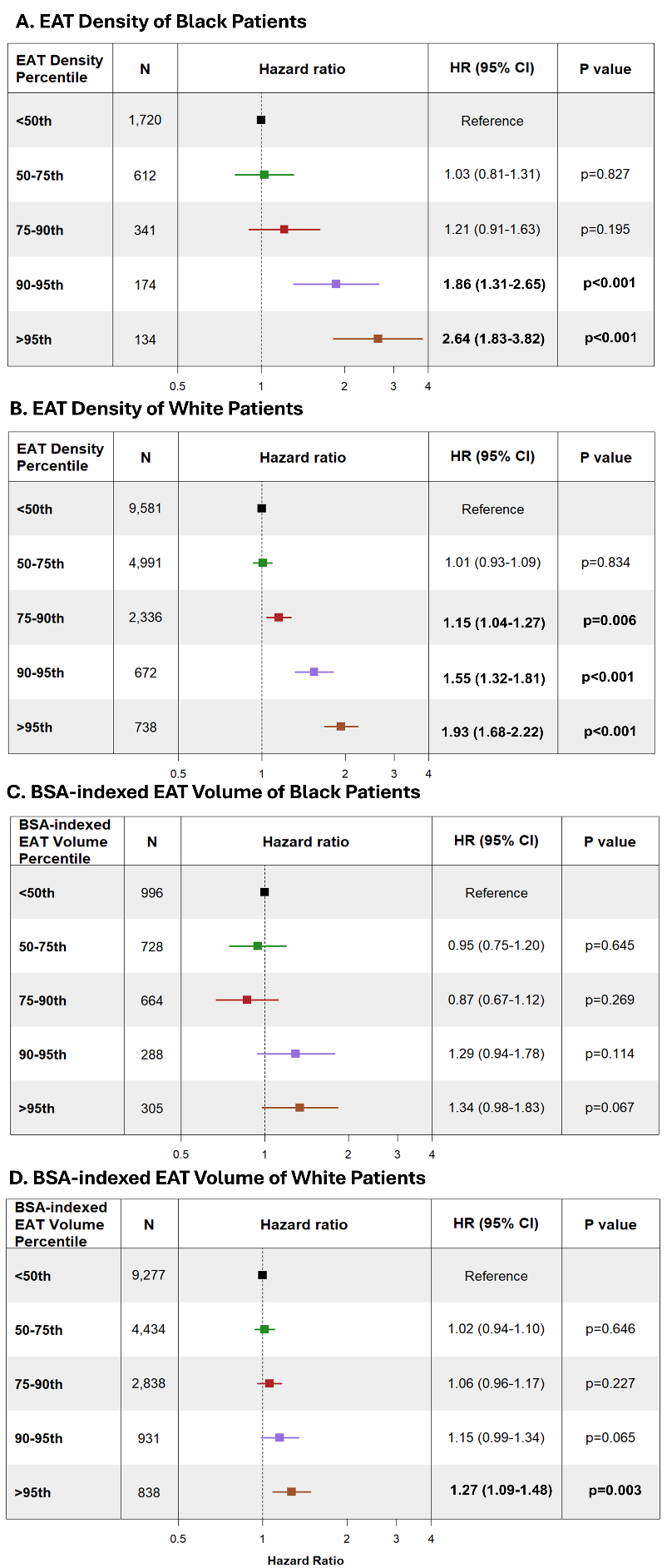

Supplemental Figure 11. Unadjusted Cox proportional hazards model of race-, sex-, and age- stratified EAT percentile groups, with patients under the 50^th^ percentile serving as the reference group. Panel A displays unadjusted hazard ratios across EAT density percentiles in Black patients. Panel B displays unadjusted hazard ratios across EAT density percentiles in White patients. Panel C displays unadjusted hazard ratios across BSA-indexed EAT percentiles in Black patients. Panel D displays unadjusted hazard ratios across BSA-indexed EAT percentiles in White patients.

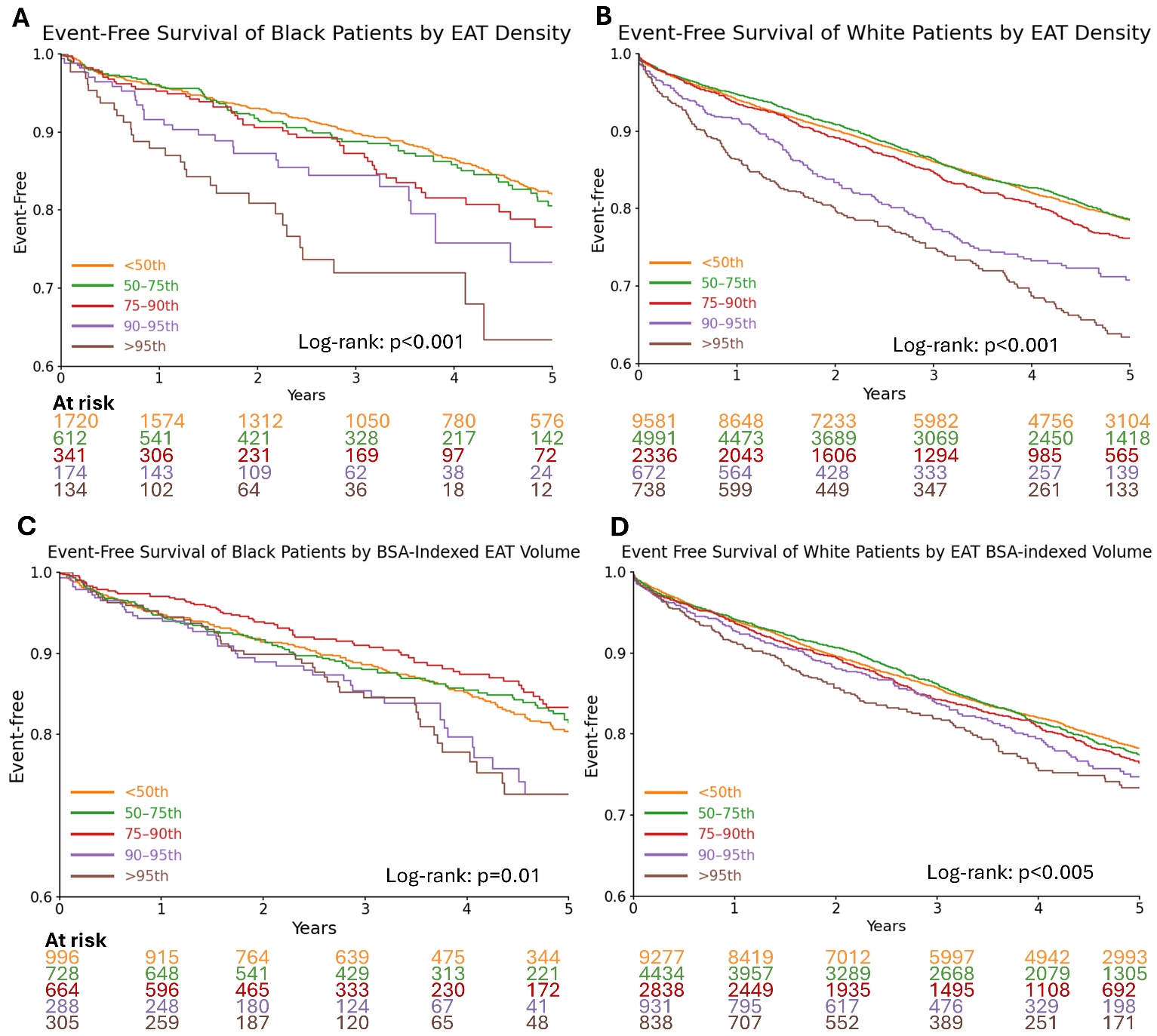
Supplemental Figure 12:

Supplemental Figure 12. Event-free Kaplan –Meier curves of EAT Density and BSA-indexed EAT volume percentile groups in Black and White patients. Percentile groups were stratified by race, sex, and age. Kaplan –Meier curves were compared using log-rank test.
